# Reprogramming of Iron and Oxygen Metabolism Across the Spectrum of Primary Aldosteronism

**DOI:** 10.64898/2026.06.09.26355256

**Authors:** Stéfanie Parisien-La Salle, Cheng-Hsuan Tsai, Andrew J Newman, Mahyar Heydarpour, Sanan Mahrokhian, Isabelle Hanna, Jenifer M Brown, Sushrut Waikar, Marwan Moussa, Anand Vaidya

## Abstract

Primary aldosteronism (PA) causes oxidative stress, myocardial injury, and increases hemoglobin, whereas aldosterone antagonists mitigate myocardial injury and lower hemoglobin. Whether these phenomena involve altered iron and oxygen metabolism is unknown. To investigate this, we leveraged human physiology studies that characterized the spectrum of PA, from subclinical to clinically overt renin-independent aldosteronism. The plasma proteome was measured to conduct aldosterone dose-response analyses to identify iron and oxygen metabolism related pathways. CYBRD1, a mediator of iron reduction and absorption, was the most abundant protein in people with overt PA, and Reactome enrichment identified 16 iron and heme-related pathways involving erythrocyte oxygen handling, heme biosynthesis, and mitochondrial respiratory electron transport. Across the continuum of PA, there were progressive decreases in mitochondrial electron transport proteins, and increases in hemoglobin subunits, heme-related proteins, and erythrocyte oxygen-handling enzymes. These proteomic changes were validated by demonstrating aldosterone dose-dependent increases in circulating hemoglobin in this cohort and an independent population-based cohort. Collectively, PA is characterized by progressive decreases in mitochondrial oxidative phosphorylation, and increases in iron absorption, heme synthesis, and oxygen-delivery, possibly reflecting a compensatory response to cellular pseudohypoxia. These findings provide a mechanistic basis for aldosterone-mediated myocardial injury and the benefits of aldosterone-directed therapy.

## Introduction

Primary aldosteronism (PA) is an adrenal disorder characterized by renin-independent aldosterone production (1). It is a common contributor to hypertension (2), highly prevalent in resistant hypertension (3–6) and obesity-related hypertension (7), and manifests across a continuum of severity that parallels risk for adverse cardiovascular and kidney outcomes (2, 8–18).

Given that hypertension is the leading modifiable risk factor for cardiovascular morbidity and mortality, a deeper understanding of the mechanisms driving PA has become increasingly important (19–23). Beyond the well described cardiovascular and renal complications associated with excess aldosterone (8, 9, 11, 12, 14, 24–26), aldosterone also influences erythrocyte physiology and oxygen handling (27–29). A recent study identified higher hemoglobin subunit beta (HBB) in patients with PA compared to those without PA (30). Conversely, clinical trials evaluating mineralocorticoid receptor (MR) antagonist therapy (with spironolactone and finerenone) have consistently demonstrated unexplained reductions in hemoglobin (31–33). From a cellular perspective, erythrocytes from patients with PA demonstrate oxidative stress-related membrane alterations mediated through the MR (27, 28) and aldosterone has been shown to shift the oxygen-hemoglobin dissociation curve, reducing hemoglobin oxygen affinity in vivo (29). Together, these observations suggest that aldosterone excess may alter red blood cell biology and heme handling. In parallel, iron has also emerged as a potential mediator of aldosterone-related tissue injury (34). Experimental studies have shown that iron restriction attenuates aldosterone/salt-induced renal injury and oxidative stress, implicating dysregulated iron handling in the downstream effects of aldosterone excess (34). In support of this concept, ferroptosis-related pathways have been implicated in aldosterone-producing adenomas (35). However, whether aldosterone excess directly reprograms iron metabolism remains unknown.

We evaluated the proteomic landscape of iron and oxygen metabolism pathways across the spectrum of PA, ranging from subclinical to clinically overt PA.

## Results

### Study Population

Baseline characteristics of the study population are shown in **Table 1**. Normotensive participants who underwent an oral sodium suppression test to quantify the degree of subclinical PA (renin-independent aldosterone production) (n=61) had a mean age of 49.4 ± 10.2 years, 49.2% were women, and most participants were White (83.6%). Mean BMI was 29.5 ± 4.4 kg/m² with mean untreated blood pressure in the normotensive range. Serum potassium was within the normal range and no participant was receiving antihypertensive medications.

**Table 1:** Baseline Characteristics.

|  | Normotensive Cohort |  |  |  | Overt Primary Aldosteronism Cohort |
| --- | --- | --- | --- | --- | --- |
|  |  | Spectrum of Subclinical Primary Aldosteronism<br>(Tertiles of Renin-Independent Aldosterone Production Following an Oral Sodium Loading Test) |  |  | Overt Renin-Independent Aldosterone Production and Volume Expansion |
|  | All<br>N=61 | T1<br>N=20 | T2<br>N=20 | T3<br>N=21 | All<br>N= 50 |
| **Age (years)** (mean, SD) | 49.4 (10.2) | 52.8 (10.33) | 49.6 (9.9) | 45.9 (9.6) | 53.7 (12.1) |
| **Female sex** (n, %) | 30 (49.2) | 13 (65.0) | 8 (40.0) | 9 (42.9) | 17 (34.0) |
| **Race** (n, %) |  |  |  |  |  |
| White | 51 (83.6) | 16 (80.0) | 19 (95.0) | 16 (76.2) | 24 (48.0) |
| Black | 5 (8.2) | 3 (15.0) | 0 (0.0) | 2 (9.5) | 17 (34.0) |
| Other | 5 (8.2) | 1 (5.0) | 1 (5.0) | 3 (14.3) | 9 (18.0) |
| **Body mass index (kg/m<sup>2</sup>)** (mean, SD) | 29.5 (4.4) | 29.4 (4.4) | 27.9 (2.8) | 31.1 (5.2) | 32.3 (6.8) |
| **Seated systolic blood pressure (mmHg)** (mean, SD) | 122.0 (7.6) | 121.1 (9.3) | 122.8 (6.2) | 121.9 (7.3) | 147.7 (20.6) |
| **Seated diastolic blood pressure (mmHg)** (mean, SD) | 77.4 (4.8) | 77.7 (5.5) | 77.8 (4.7) | 76.6 (4.2) | 89.5 (12.2) |
| **Serum potassium (mmol/L)** (mean, SD) | 4.3 (0.3) | 4.3 (0.3) | 4.3 (0.3) | 4.3 (0.4) | 3.4 (0.6) |
| **Urinary Potassium-to-Sodium ratio**, (mean, SD) | 0.18 (0.1) | 0.16 (0.05) | 0.18 (0.1) | 0.20 (0.1) | - |
| **Estimated Glomerular Filtration Rate (ml/min)** (mean, SD) | 89.3 (12.41) | 91.0 (12.4) | 89.3 (12.1) | 87.6 (13.1) | 67.2 (16.9) |
| **Seated Measurements\*** (med, IQR) |  |  |  |  |  |
| Plasma Renin Activity (ng/mL/hr) | 0.4 (0.2 - 0.6) | 0.2 (0.1 - 0.33) | 0.4 (0.2 - 0.53) | 0.6 (0.3 - 1) | 0.3 (0.1-0.6) |
| Aldosterone (ng/dL) | 9.1 (7.4-16.6) | 7.1 (5.8-8.5) | 9.6 (8.3-12.3) | 21.6 (14.4 -29.3) | 23 (15.7-37.0) |
| **Supine Measurements\*** (med, IQR) |  |  |  |  |  |
| Plasma Renin Activity (ng/mL/hr) | 0.2 (0.1 - 0.4) | 0.1 (0.1 - 0.2) | 0.2 (0.1 - 0.3) | 0.3 (0.1 - 0.6) | - |
| Aldosterone (ng/dL) | 7.6 (5.9 – 11.7) | 5.2 (4.3 - 5.7) | 7.5 (7.2 – 9.0) | 13.7 (11.7 - 24.3) | - |
| **Number of hypertensive medications** (mean, SD) | 0 (0.0) | 0 (0.0) | 0 (0.0) | 0 (0.0) | 3 (1.0) |

As expected, participants with clinically overt PA (N=50) had severe renin-independent aldosteronism, were older (53.7 ± 12.1 years), had higher BMI (32.3 ± 6.8 kg/m²), and higher blood pressure, despite treatment with on average 3 anti-hypertensive medications. Participants with PA were also predominantly male, and had lower serum potassium and estimated glomerular filtration rate when compared to the normotensive participants.

### Enrichment of Iron and Heme Biology Pathways When Categorically Comparing Overt PA to Normotensives

As previously demonstrated, 903 proteins in the peripheral circulation were differentially abundant in Overt PA when compared with individuals with normotension (**Supplemental Figure 1**) (36). To characterize the iron, heme, and oxygen metabolic processes distinguishing PA from normotension, Reactome pathway enrichment analysis was performed on differentially expressed proteins and applied keyword-based filtering to focus on heme metabolism, iron handling, erythrocyte biology and mitochondrial energy metabolism. This approach identified 16 significantly enriched pathways (p < 0.10) (**Figure 1A**), with the tricarboxylic acid (TCA) cycle and respiratory electron transport pathway showing the highest protein ratio, followed by cytoprotection by HMOX1 and erythropoietin signaling. Further enriched pathways included heme biosynthesis, heme degradation, heme scavenging, porphyrin metabolism, iron uptake and transport, O2/CO2 exchange in erythrocytes and mitochondrial iron-sulfur cluster biogenesis.

**Figure 1:**
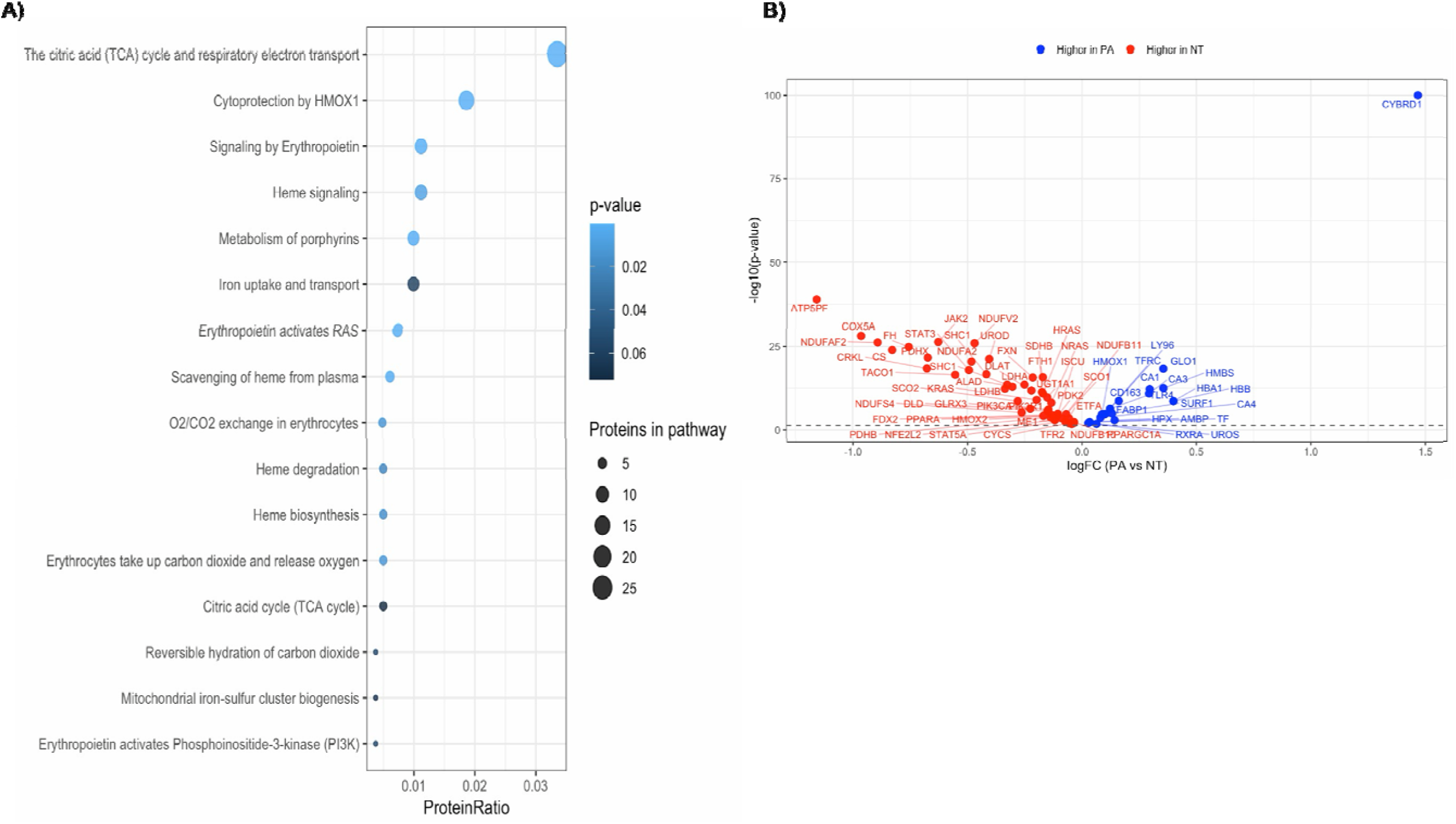
Reactome Pathway Enrichment and Differential Abundance of Iron and Heme-Related Proteins in Primary Aldosteronism. A) Reactome pathway enrichment of 903 differentially abundant proteins filtered for iron and heme-related terms, yielding 16 pathways (p<0.1). Dot size reflects the number of proteins per pathway and color indicates p-value. B) Volcano plot of proteins belonging to these 16 iron and heme-related pathways. **Blue** dots indicate proteins higher in PA. **Red** dots indicate proteins higher in normotensive (NT) participants.

A volcano plot reveals the landscape of differentially abundant proteins driving these pathways (**Figure 1B**). CYBRD1, a duodenal cytochrome b reductase critical for dietary iron reduction, emerged as the most significantly abundant protein in PA (logFC = 1.47, adj. p-value = 1.36×10⁻&#x25A1;⁻&#x25A1;). Additional proteins significantly more abundant in PA included carbonic anhydrases CA1, CA3 and CA4, consistent with enhanced CO2/O2 exchange capacity, hemoglobin subunits HBB/HBA1, heme biosynthesis enzymes HMBS and UROS, the heme scavenging protein HPX and HMOX1 and iron transport protein TF. In contrast, multiple TCA cycle, pyruvate dehydrogenase complex and mitochondrial electron transport proteins were broadly lower in PA. Iron storage, such as ferritin heavy chain (FTH1) was also less abundant in PA. To visualize the directionality of individual protein contributions across these pathways, we constructed a protein-by-pathway heatmap (**Figure 2**). Proteins associated with TCA cycle, mitochondrial electron transport chain and erythropoietin signaling were generally less abundant in PA relative to normotensive people, whereas proteins mapping to erythrocyte-specific pathways, including heme biosynthesis, heme scavenging and O2/CO2 exchange, were predominantly more abundant in PA. Collectively, these findings suggested that PA is characterized by a proteomic signature consistent with impaired mitochondrial oxidative phosphorylation alongside increased erythrocyte activity and heme and iron turnover.

**Figure 2:**
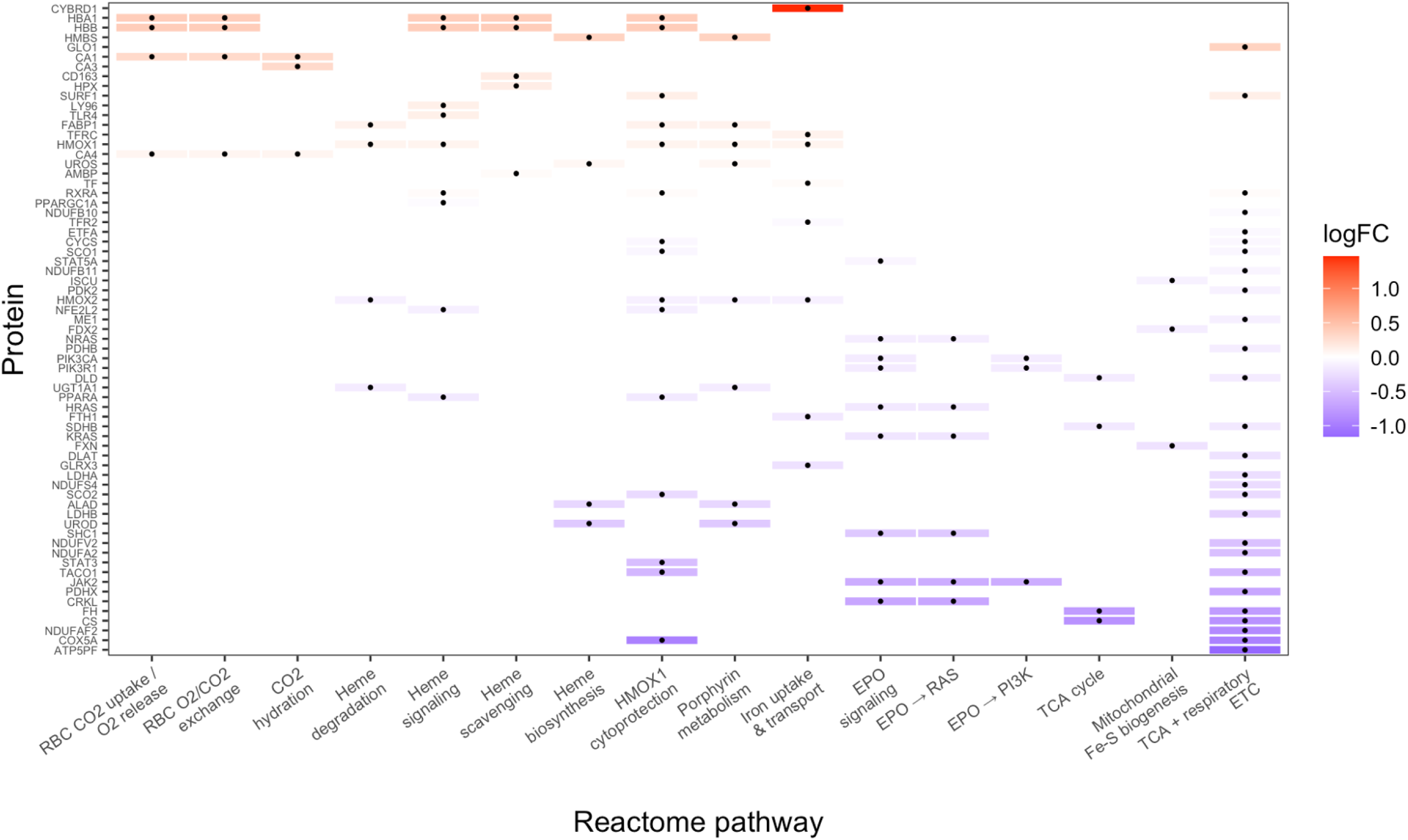
Protein-by-Pathway Heatmap of Iron and Heme-Related Proteins in Primary Aldosteronism. Each row represents a differentially abundant protein and each column a Reactome iron or heme-related pathway. Cell color reflects the log fold change (logFC) of each protein in PA versus normotensive participants.

### Proteomic Trends Consistent with Iron and Heme Biology Re-Programming Across the Spectrum of PA

To explore whether proteins within the enriched heme, iron and oxygen metabolism-related pathways showed progressive changes across the PA continuum, we examined their abundance across tertiles of subclinical PA (SubPA) in normotensive people and among those with overt PA (**Figure 3A-B**). There were 14 iron metabolism-related proteins demonstrating monotonic trends across the PA continuum. Proteins displaying an upward trend across the continuum (SubPA1-SubPA2-SubPA3-Overt PA) were enriched for hemoglobin subunits (HBA1/HBB), erythrocyte metabolism (GLO1, CA1, CA3), heme biosynthesis (HMBS, UROS) and heme scavenging (HPX, AMBP). These proteins largely overlapped with those showing the highest relative abundance in the categorical comparisons between overt PA and non-PA, further supporting the effect of aldosterone excess on iron and heme regulation. In contrast, proteins demonstrating a downward trend spanned mitochondrial electron transport (CYCS, ETFA) and hepatic heme catabolite clearance (UGT1A1) (**Figure 3A-B**).

**Figure 3:**
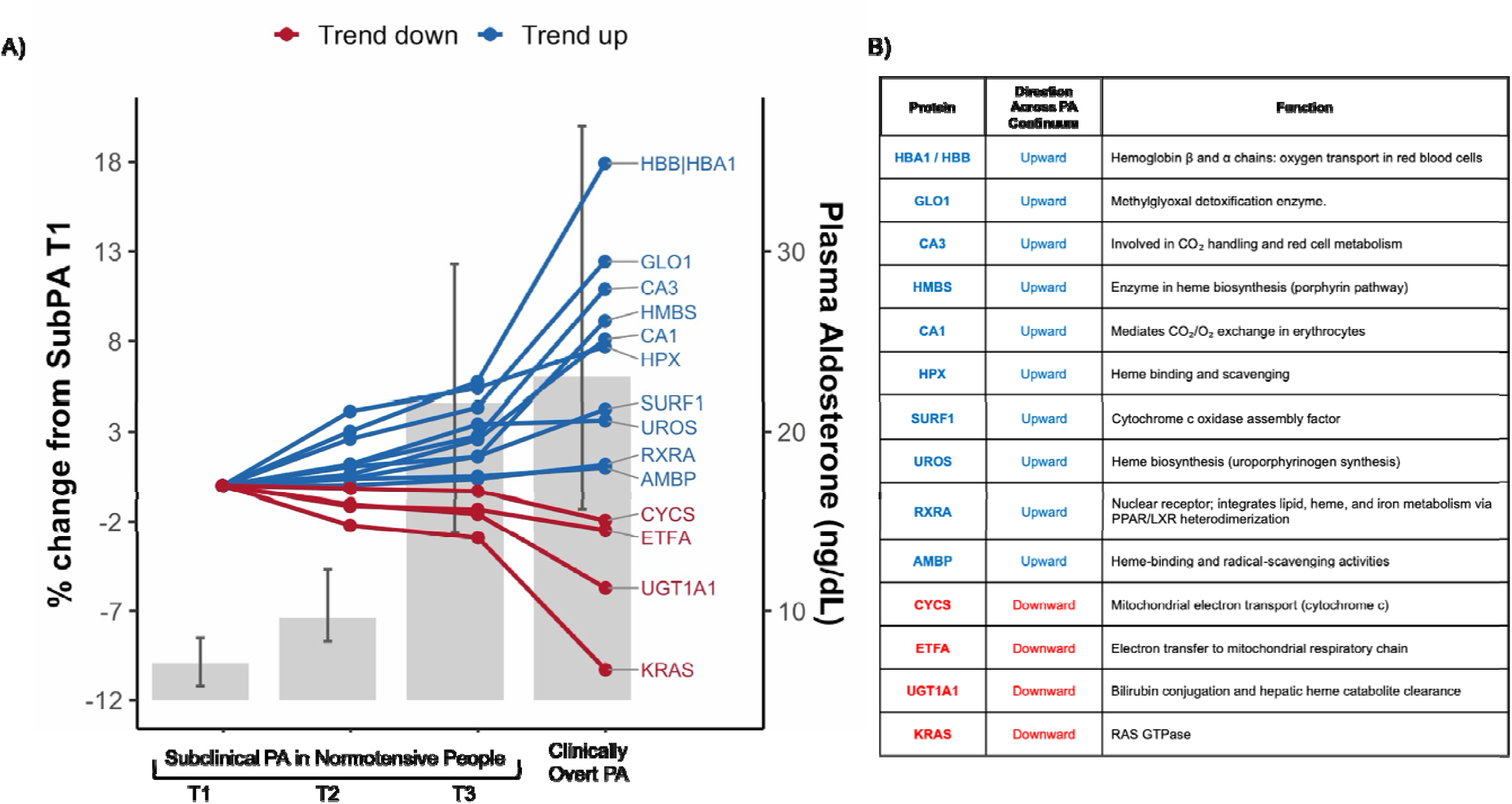
Iron and Heme-Related Plasma Proteins Across the Primary Aldosteronism Continuum. A) Percent change in plasma protein levels relative to subclinical PA tertile 1 (T1) across increasing aldosterone exposure. Blue and red lines indicate proteins with upward and downward trends, respectively. Gray bars represent mean plasma aldosterone (right y-axis). B) Table of the trending proteins, highlighting enrichment for heme biosynthesis, oxygen transport, and mitochondrial electron transport functions.

### Comprehensive Integration of Iron and Heme Metabolism Changes in PA

**Figure 4** illustrates the integration of all the observed proteomic changes and directions in iron and heme-related proteins in PA. Collectively, these changes are consistent with a systemic shift away from mitochondrial oxidative phosphorylation and toward increased intestinal iron absorption, preferential iron transport over storage, and enhanced heme synthesis and recycling.

**Figure 4:**
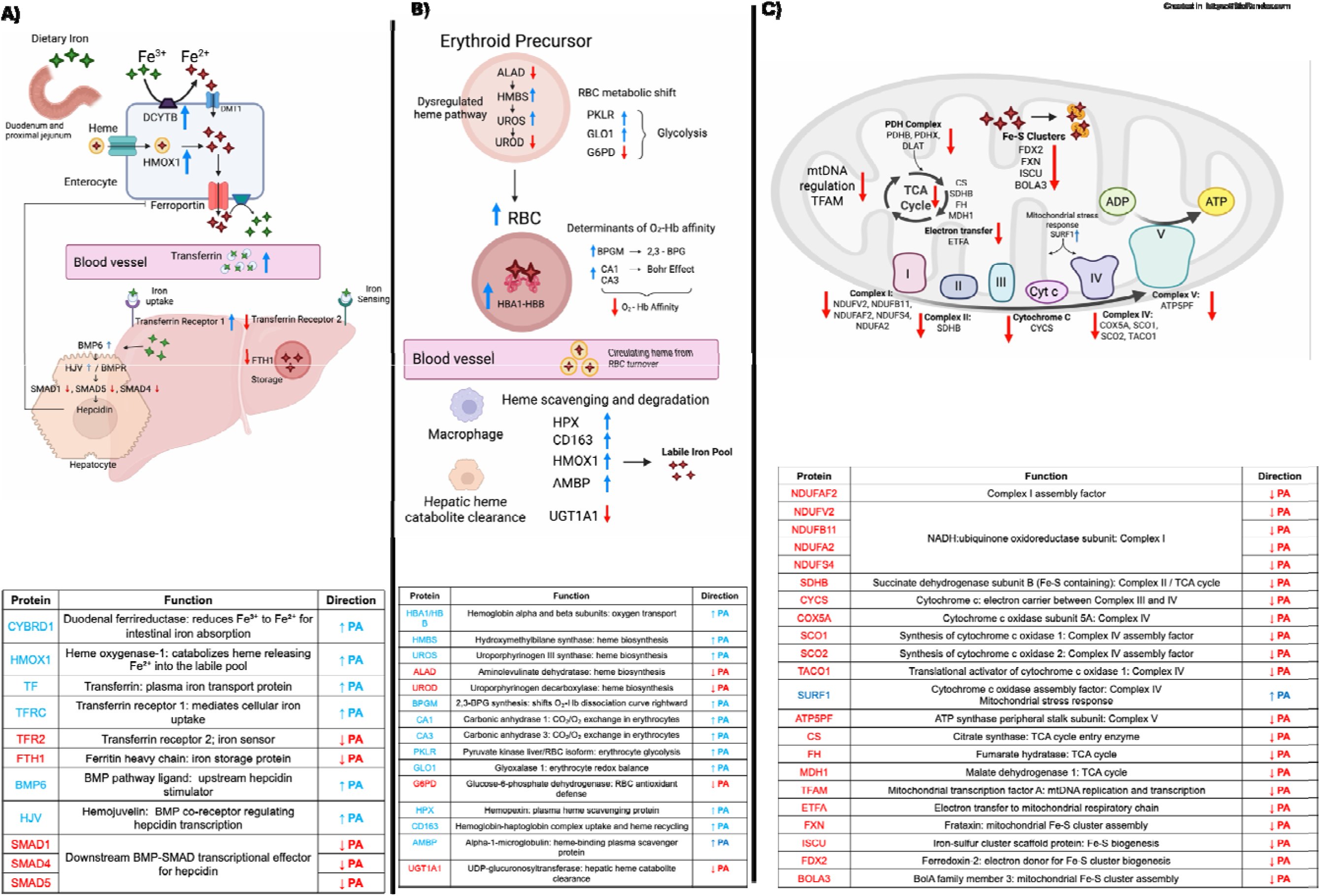
Comprehensive Integration of Iron and Heme Metabolism Changes in PA. **Panel A** illustrates dysregulated intestinal iron absorption and impaired hepatic hepcidin regulation. CYBRD1 (DCYTB) was the most differentially abundant protein in PA, indicating markedly enhanced ferrireductase activity in the duodenum, facilitating iron absorption. HMOX1 elevation indicates increased catabolism of heme, releasing additional Fe²⁺ for export via ferroportin. Downstream, transferrin and transferrin receptor 1 were elevated, indicating increased iron transport and cellular uptake, while transferrin receptor 2 (an iron sensing protein), and ferritin heavy chain (FTH1) were reduced, suggesting preferential iron mobilization over storage. Within the hepatocyte, BMP6 and HJV were elevated, yet SMAD1, SMAD4 and SMAD5 were all significantly lower in PA, indicating disruption of the BMP-SMAD transduction cascade required for hepcidin transcription. This suggests that hepcidin output is possibly insufficient to restrain iron import. **Panel B** shows that elevated iron availability in PA supports erythroid expansion. Heme biosynthesis was dysregulated, with HMBS and UROS more abundant and ALAD and UROD less abundant in PA, suggesting altered flux through the porphyrin pathway. Hemoglobin subunits HBA1/ HBB were among the most significantly elevated proteins, consistent with increased red blood cell production. Elevation of BPGM and CA1/CA3 indicates enhanced 2,3-BPG synthesis and carbonic anhydrase activity, which together shift the oxygen-hemoglobin dissociation curve rightward and reduce O₂-hemoglobin affinity, leading to increased oxygen delivery. PKLR and GLO1 were elevated while G6PD was reduced, consistent with a glycolytic shift in RBC metabolism. Finally, elevation of HPX, CD163, HMOX1 and AMBP indicate heme scavenging from circulating RBC turnover, sustaining a labile iron pool that may further be used for erythropoiesis. Reduced UGT1A1 suggests impaired hepatic heme catabolite clearance, consistent with iron being recycled toward erythropoiesis rather than excreted. **Panel C** shows that increased iron demand in PA comes at the expense of mitochondrial function. Multiple electron transport chain subunits were lower in PA, from Complex I (NDUFV2, NDUFB11, NDUFAF2, NDUFS4, NDUFA2), Complex II (SDHB), Complex IV (COX5A, SCO1, SCO2, TACO1) and Complex V (ATP5PF), with cytochrome c (CYCS). Interestingly, SURF1, a Complex IV assembly factor was elevated in PA. TCA cycle enzymes citrate synthase (CS), fumarate hydratase (FH) and malate dehydrogenase (MDH1) were reduced, as well as components of the pyruvate dehydrogenase complex (PDHB, PDHX, DLAT). Mitochondrial transcription factor A (TFAM), which regulates mtDNA replication and transcription, was also lower in PA. Proteins required for mitochondrial iron-sulfur cluster assembly (FDX2, FXN, ISCU and BOLA3) were all reduced, suggesting that impaired Fe-S biogenesis may represent a link between iron redistribution toward erythropoiesis and respiratory chain dysfunction. Collectively, these proteomic changes are consistent with a systemic shift in PA toward increased intestinal iron absorption, enhanced circulating iron transport over storage and preferential heme synthesis and recycling at the expense of mitochondrial oxidative phosphorylation capacity. **Blue** arrows indicate proteins more abundant in PA. **Red** arrows indicate proteins less abundant in PA.

### Proteomic Analyses After Adjustment for Clinical Covariates

To assess whether the proteomic findings were related specifically to the continuum of PA pathophysiology, all analyses were repeated after adjustment for age, sex, body mass index, systolic blood pressure, serum potassium, and glomerular filtration rate to demonstrate that the observed findings were independent of these modifying factors. Differential abundance testing identified 526 proteins in the adjusted model (**Supplemental Figure 2**). Although a subset of proteins no longer met significance criteria after co-variate adjustments, Reactome pathway enrichment identified 11 iron and heme-related pathways as significantly enriched, with a consistent pattern of directionality at the individual protein level (**Supplemental Figure 3-4**). Aldosterone dose-dependent trends across the PA continuum also remained largely unchanged after adjustment (**Supplemental Figure 5**).

### Clinical Correlation with Blood Count Parameters

To validate these proteomic discoveries, aldosterone dose-response relationships with circulating hemoglobin, hematocrit and red blood cell count were examined in three different approaches: 1) overall differences between the normotensive cohort and the overt PA cohort; 2) differences across normotensive tertiles of SubPA and overt PA; and 3) stratifying patients with overt PA based on their ultimate subtype determined by adrenal venous sampling (**Figure 5**). Across all analyses, higher degrees of renin-independent aldosterone production were associated with progressive increases in all blood count parameters. These increases were most statistically significant for red blood cell count, while hemoglobin and hematocrit demonstrated similar upward trends that did not consistently reach statistical significance.

**Figure 5:**
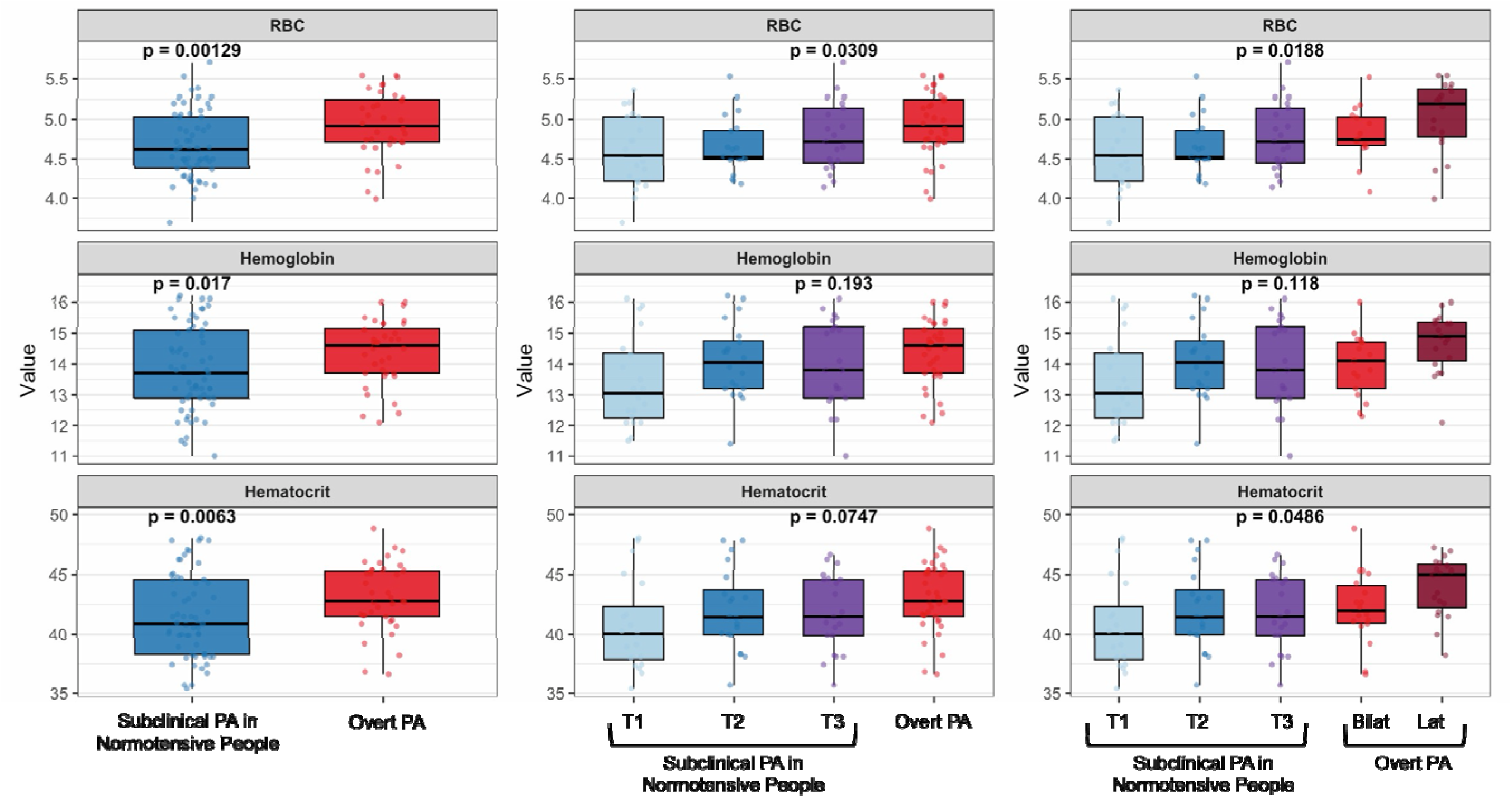
Red Blood Cell Parameters Across the Primary Aldosteronism Continuum. Boxplots of red blood cell count (RBC), hemoglobin, and hematocrit across three comparisons: subclinical PA in normotensive individuals versus overt PA (left column), subclinical PA tertiles T1-T3 versus overt PA (middle column) and subclinical PA tertiles versus overt PA stratified by bilateral or lateral disease (right column). Two-group comparisons were assessed using sex-adjusted linear models and trends across aldosterone categories were evaluated using sex-adjusted rank-based trend tests. Individual data points are overlaid on each boxplot.

### External Validation in Population-Based Cohort

In an independent population-based cohort of 5,713 patients with hypertension, hemoglobin levels were progressively higher in parallel with greater renin-independent aldosterone production. Median plasma renin activity was 0.3 [0.1, 0.6] ng/mL/h and plasma aldosterone concentration ranged from 8.1 [5.1, 10.6] ng/dL in Q1 to 44.9 [37.2, 61.2] ng/dL in Q4, while hemoglobin increased from 12.3 ± 2.4 g/dL to 13.3 ± 2.0 g/dL across the same quartiles (**Supplemental Figure 6 and Supplemental Table 2**). This association remained statistically significant after adjustment for age, sex, estimated glomerular filtration rate, serum potassium, and antihypertensive medication burden (adjusted P for trend <0.001), supporting a positive relationship between higher renin-independent aldosterone exposure and hemoglobin in a real-world hypertensive cohort who underwent PA testing.

## Discussion

We found extensive and progressive reprogramming of iron and heme metabolism across the continuum of PA. These changes were consistent with a coordinated redirection of iron away from mitochondrial oxidative phosphorylation and toward increased intestinal iron absorption, preferential iron transport over storage, and enhanced heme synthesis and recycling. Notably, key proteins within this signature showed progressive, aldosterone dose-dependent changes across the PA continuum, implicating dysregulated iron metabolism as an early and persistent manifestation of the disorder. The iron metabolic reprogramming we describe may have broader consequences for iron-dependent organs, such as the myocardium, where impaired mitochondrial oxidative phosphorylation and iron-sulfur cluster biogenesis could provide a novel mechanistic basis for the cardiovascular disease seen in PA (8, 16, 26, 37–39).

While the cross-sectional nature of our study cannot confirm causality, we propose the following model: aldosterone-mediated impairment of cellular iron uptake depletes tissues of the iron required for mitochondrial Fe- S cluster biogenesis and impairs oxidative phosphorylation, thus promoting cellular pseudohypoxia and tissue injury. The resulting accumulation of iron in the labile pool, is then redirected toward EPO-independent erythroid expansion (40) increasing hemoglobin mass and shifting the oxygen-hemoglobin dissociation curve rightward, as a compensatory attempt to deliver more oxygen to metabolically compromised tissues. An alternate hypothesis is that aldosterone may directly drive erythroid expansion (41) channeling iron toward heme at the expense of mitochondrial Fe-S cluster biogenesis and oxidative phosphorylation. Collectively, these findings may simultaneously provide a mechanism for aldosterone-mediated tissue injury, the reactive response to this pathology, and the mechanism by which aldosterone-directed therapy may prevent tissue injury.

A comprehensive assessment of proteomic data across the continuum of PA revealed a broad reduction in proteins involved in electron transport chain subunits across Complexes I, II, IV, and V, tricarboxylic acid (TCA) cycle enzymes, pyruvate dehydrogenase (PDH) enzymes and mitochondrial DNA transcription. Proteins required for mitochondrial iron-sulfur (Fe-S) cluster assembly were also reduced (**Figure 4C**). Fe-S clusters are essential cofactors serving as critical mediators of electron transfer within the mitochondrial respiratory chain (42). Fe-S cluster deficiency impairs respiratory complexes I-III directly (as Fe-S-containing enzymes) and Complex IV indirectly (via impaired translation of its subunits, which depend on Fe-S cluster assembly factors) (42, 43). Fe-S clusters are also essential for TCA cycle function and for PDH activity (42). Our findings are consistent with Hung et al.(44) and Tsai et al.(45), who demonstrated that aldosterone can directly suppress cardiac mitochondrial DNA copy number, COX IV, SOD2 and ATP production through MR-dependent oxidative stress pathways, with effects reversed by eplerenone in both cell and animal models (44, 45). Also, a myocardial proteomics study using the SomaLogic platform in heart failure found lower abundance of proteins related to oxidative phosphorylation, with these changes being more pronounced in a proteomic subgroup enriched for severe obesity (46), a phenotype increasingly recognized to overlap with pathogenic aldosteronism (7).

A growing body of evidence implicates aldosterone in myocardial iron deficiency and its downstream consequences for cardiac function (38, 47, 48). Aldosterone has been proposed to promote myocardial iron deficiency through downregulation of transferrin receptor 1 on cardiomyocytes, potentially reducing cellular iron uptake in the heart and thereby limiting the iron availability required for Fe-S cluster biogenesis and mitochondrial function (38). Myocardial iron deficiency has been directly linked to suppressed respiratory chain activity, impaired ATP production and adverse cardiac remodeling in heart failure (49). Together, these findings raise the possibility that aldosterone-mediated tissue iron deficiency may contribute to PA-associated organ injury, and that targeting this mechanism through aldosterone-directed therapy may attenuate its consequences.

Another notable finding in patients with PA was the shift in iron and heme metabolism directed toward erythroid expansion, characterized by enhanced intestinal iron absorption, increased plasma iron transport over storage, active heme recycling, dysregulated heme biosynthesis, ultimately resulting in higher levels of hemoglobin subunits (**Figure 4A & 4B**). HBA1/HBB levels were positively associated with aldosterone dose- dependent trends across the PA continuum, with progressively higher abundance paralleling disease severity. Importantly, these findings were confirmed by corresponding increases in complete blood count parameters, with dose-dependent elevations in red blood cell count, hemoglobin and hematocrit in both our primary cohort and the large validation cohort of over 5000 hypertensive patients. As mentioned previously, Tsai et al. reported HBB as one of six peptides that could distinguish PA from essential hypertension, with higher levels in PA that declined following aldosterone-directed therapy (30). In a study of vascular inflammation and immune cell phenotypes in PA, van der Heijden et al. observed higher HBA1 expression in monocytes from patients with PA (50). Conversely, in FIDELITY, a pooled analysis of the FIDELIO-DKD and FIGARO-DKD clinical trials (n ≍13,000) evaluating finerenone in patients with CKD and type 2 diabetes, hemoglobin levels declined in the first 4 months with finerenone compared with placebo in patients without baseline anemia (32). Similarly, in Aldo-DHF, a randomized, placebo-controlled trial of spironolactone 25 mg daily in 422 patients with HFpEF, a decrease in hemoglobin was observed in the spironolactone group at 12 months, a finding the investigators described as unexplained (31).

Finally, In FINEARTS-HF, hemoglobin declined more with finerenone than with placebo during follow-up (33). These clinical trial findings provide causal evidence that pharmacologic antagonism of aldosterone reduces hemoglobin, thereby lending support to our current findings that indicate shunting of iron metabolism towards increased erythroid expansion in parallel with pathologic aldosteronism.

Although we could not control for obstructive sleep apnea, a condition that may contribute to increased hemoglobin levels (51), the observed trend persisted when analyses were restricted to normotensive participants, who are not considered to be at high risk for sleep apnea. Moreover, our pathway analysis suggested that the erythroid process may possibly be EPO-independent, as reflected by the lower abundance of proteins involved in EPO-related pathways (**Figure 2**).

Beyond PA, hemoglobin has also been reported to correlate with blood pressure (52–55). Although there are several mechanistic reasons to potentially explain the link between blood pressure and hemoglobin, perhaps the most underappreciated is that PA is grossly underdiagnosed in people with hypertension. Importantly, our observations remained robust after multiple adjustments including blood pressure, suggesting that the erythroid proteomic signature reflects aldosterone-driven biology beyond the effects of blood pressure. Our data also revealed changes in heme biosynthesis and red blood cell physiology. The elevation of BPGM and carbonic anhydrases CA1 and CA3, consistent with a rightward shift of the oxygen-hemoglobin dissociation curve and enhanced oxygen delivery, may reflect both the expanded erythroid mass and a direct effect of aldosterone on hemoglobin oxygen affinity, as previously demonstrated in vivo (29). Heme scavenging proteins were also more abundant, with elevation of HPX, CD163, AMBP and HMOX1. These proteins capture and catabolize free heme, liberating Fe²⁺ that re-enters the labile iron pool to sustain ongoing erythropoiesis.

While this study provides a robust proteomic view of iron and heme metabolism in PA, some limitations should be noted. First, proteomic findings in the peripheral blood may not fully reflect tissue-specific processes in key iron-regulating organs such as the liver, kidney and heart. Nevertheless, the alignment of our findings with prior functional assessments of tissue oxidative phosphorylation supports the biological relevance of the circulating signature (44–46). Moreover, using the same SomaLogic platform employed in our study, Zampino et al. demonstrated that plasma mitochondrial proteins correlate with in vivo skeletal muscle oxidative phosphorylation capacity assessed by phosphorous magnetic resonance spectroscopy, the gold-standard non- invasive measure of tissue mitochondrial function (56). Second, important components of the iron regulatory axis, including a fuller characterization of hepcidin signaling, remain incompletely captured by our proteomic dataset and warrant further investigation. Third, we did not measure serum iron, total iron-binding capacity or ferritin, and therefore our conclusions are mostly based on the proteomic data and blood count parameters. Finally, as a cross-sectional analysis, our study captures a single proteomic snapshot and cannot establish causality of the observed iron changes. However, the implications of our study involving iron, heme, and cellular respiration phenomena may explain the deleterious effects of PA on several tissues and organs, such as the myocardium, where aldosterone-directed therapies such as mineralocorticoid receptor antagonists have demonstrated overwhelming clinical benefits (57, 58).

In conclusion, we show that the spectrum of primary aldosteronism is characterized by a progressive and coordinated redirection of iron away from mitochondrial Fe-S cluster biogenesis and oxidative phosphorylation, potentially contributing to cellular pseudohypoxia and tissue injury, and toward erythroid expansion with compensatory increases in gastrointestinal iron absorption and tissue iron delivery. By linking aldosterone excess to altered iron handling, heme metabolism, and impaired mitochondrial oxidative phosphorylation, these findings offer a novel mechanistic basis for aldosterone-mediated tissue injury and provide explanations for the beneficial effects of aldosterone-directed therapy.

## METHODS

### Sex as a biological variable

Both female and male participants were included in this study. Sex was considered as a biological variable in the study design. Accordingly, sex was included as a covariate in adjusted proteomic analyses and in analyses of blood count parameters.

### Overall Study Rationale and Design

In a previous large-scale, unbiased, proteomic analysis, we reported differences in the plasma proteome between people with and without PA (36). A key observation from that analysis was that many differentially abundant proteins in PA belonged to hemoglobin and iron-related biological pathways. Given the known role of the renin-angiotensin-aldosterone system in erythropoiesis (41, 59, 60), and prior reports of increased hemoglobin abundance in PA (30), we hypothesized that the severity spectrum of PA might be accompanied by parallel changes in iron and heme metabolism. In this study, we conducted targeted secondary analyses of proteomic changes across the PA continuum related to iron metabolism and hemoglobin-related pathways.

The analyses involved investigation of the peripheral plasma proteome of participants with clinically overt PA compared to that of normotensive participants who had undergone an oral sodium loading test (OSLT) to evaluate the degree of renin-independent aldosterone production (subclinical PA). The proteomic signature associated with PA was evaluated as a binary construct, PA vs no PA, and also as an ordinal dose-response construct, using monotonic trends across the PA continuum (subclinical to overt), defined by the magnitude of renin-independent aldosterone production. Pathway enrichment was then performed to identify iron and erythroid-related biological processes.

### Study Population

Participants were originally recruited for two prospective study protocols: (1) a well-described community-based cohort of normotensive individuals with risk factors for developing incident hypertension who underwent deep phenotyping procedures to characterize the presence and degree of subclinical PA (n=61) (36, 61–63) and (2) a prospective registry of patients with clinically overt PA (n=50).

### Phenotyping Maneuvers

As previously described, normotensive participants underwent an OSLT to assess the degree of pathologic, renin-independent aldosterone production and were subsequently stratified into unbiased tertiles based on plasma aldosterone concentrations during the sodium load. The OSLT involved consuming >200 mmol/d of sodium for 5-7 days, following which the combination of supine posture and sodium loading was used to induce a physiologic nadir in aldosterone production to characterize the spectrum of subclinical PA in normotensive people (61). Patients with clinically overt PA were already in a volume expanded state and renin- independent aldosteronism state owing to their underlying condition (**Table 1**).

### Targeted Iron and Erythroid Pathway Analysis

The comparison of plasma proteomic differences between individuals with overt PA versus normotensive individuals had previously yielded 903 differentially abundant proteins (36). The most statistically significant protein in participants with PA was CYBRD1, a ferric reductase involved in iron homeostasis and absorption.

Pathway enrichment analysis on these 903 differentially abundant proteins was conducted using Reactome (64) which identified 392 unique biological pathways. To investigate the role of iron and heme metabolism in PA, these 392 pathways were filtered using the keywords “iron,” “heme,” “erythro,” “porphyrin,” “HMOX1,” “TCA,” “carbon dioxide,” and “oxygen,” yielding 16 pathways related to iron and heme metabolism. To capture a broader set of potentially relevant pathways, a more inclusive enrichment cutoff of p < 0.10 was used. Because this keyword- filtered analysis was intended to identify biologically coherent iron/heme signatures rather than to generate a stand-alone diagnostic biomarker panel, enriched pathways were carried forward only when supported by consistent protein-level directionality and/or aldosterone dose-response trends. Proteins within these pathways were examined for direction of differential abundance in PA versus normotensive controls, and for monotonic trends across the PA severity continuum, from subclinical PA in normotensive individuals to overt PA in clinical patients.

### Clinical Correlates of Proteomic Findings

Complete blood count parameters reflecting commonly available, but crude, markers of iron and heme status (hemoglobin, hematocrit, and red blood cell count) were analyzed as clinical correlates of the proteomic findings across the PA continuum.

### External Validation in a Population-Based Cohort

To determine whether the aldosterone-associated complete blood count changes observed in the discovery cohort were detectable in a larger real-world clinical population, we analyzed an independent population-based cohort of people with hypertension. The validation cohort was a retrospective multicenter cohort from the National Taiwan University Hospital health systems. Adult patients aged 18 years or older who underwent biochemical evaluation for hypertension were included if they had available plasma aldosterone concentration, plasma renin activity, and hemoglobin measurements. To focus on patients with renin-independent aldosterone production, we analyzed patients (n=5,713) with a low plasma renin activity (≤1 ng/mL/hr) who also had aldosterone measurements. Hemoglobin concentrations were then compared across quartiles of aldosterone to investigate changes in hemoglobin as a function of the continuum of renin-independent aldosteronism.

### Laboratory Assays

Participants had plasma aldosterone concentration (PAC) and plasma renin activity (PRA) measured as previously described (61, 65, 66).

### Plasma Proteomics

Proteomic measurements were performed using the SomaScan® assay, an aptamer-based platform that quantifies proteins using chemically modified single-stranded DNA aptamers called SOMAmers (Slow Off-rate Modifed Aptamer) (67). Detailed methods of the SomaScan® assay have been previously described (36, 63, 68). Proteomic profiles were characterized using the 7k SomaScan assay v4.1 for the normotensive participants and the SomaScan® 11K Assay v5.0 (SomaLogic, Inc.; Boulder, CO, USA) for overt PA participants. 1500 proteins were measured to reflect cardiovascular, metabolic, and inflammation profiles and are shown in **Supplemental Table 1**. The SomaLogic normalization procedure, including adaptive normalization by maximum likelihood, was used for the SomaScan® data for all analyses. Data from the 11K SomaScan assay were bridged to the 7K format using the SomaDataIO R package for cross-platform comparability (69). Data were processed in R v4.3.2 using the SomaDataIO package for loading raw proteomic data. Normality of protein abundance distributions was evaluated using visual inspection (i.e., histograms, QQ plots), and log-transformation (log_10_) was applied to approximate normality and stabilize variance.

### Statistical Analysis

Categorical variables were reported as percentages, normally distributed variables were reported as mean ± standard deviation, and non-normally distributed variables were reported as median (Q1-Q3 interquartile range). Differential abundance of plasma proteins between normotensive participants and those with overt PA was assessed using the limma package in R, which applies linear modeling with empirical Bayes moderation to improve variance estimation. Protein-level P values were adjusted for multiple testing using the Benjamini- Hochberg false discovery rate approach; adjusted P values are reported for differential abundance analyses.

Pathway enrichment analysis on these 903 differentially abundant proteins was conducted using Reactome (64). Proteins within pathways of interest were visualized using a volcano plot. Proteomic trend analyses were performed using linear regression across 4 categories of the PA continuum: unbiased tertiles of subclinical PA in normotensive people (SubPA: termed T1, T2, T3) and the peripheral circulation of participants with overt PA. For trend analyses, PA-continuum category was modeled as an ordinal variable to test for monotonic changes in protein abundance across increasing aldosterone exposure. All proteomic analyses were first performed without covariate adjustment. To assess the robustness of findings, analyses were repeated after adjustment for age, sex, body mass index, systolic blood pressure, serum potassium and estimated glomerular filtration rate.

For the complete blood count analysis, the 2-group comparison between normotensives and PA was assessed using a sex-adjusted linear model, whereas trends in hemoglobin, hematocrit and red blood cell count across aldosterone categories were evaluated using sex-adjusted rank-based trend tests. Sex adjustment was prespecified because hemoglobin, hematocrit, and red blood cell count differ substantially by sex and because the overt PA cohort had a different sex distribution than the normotensive cohort.

In the validation cohort, the association between increasing aldosterone quartiles and hemoglobin was evaluated using trend analyses. Multivariable linear regression models were used to estimate the adjusted P for trend after accounting for potential confounders, including age, sex, estimated glomerular filtration rate, serum potassium, and antihypertensive medication burden. Quartile was modeled as an ordinal exposure for the primary trend test, and hemoglobin was modeled as a continuous outcome. Statistical significance was defined as a two-sided P value <0.05.

### Study Approval

All participants provided written informed consent, and the study protocol was approved by the Mass General Brigham Institutional Review Board.

## Data Availability

The data supporting the findings of this study are available from the corresponding author upon reasonable request.

## Author Contribution

SPL and AV designed the research studies. IH, SM, AJN, SW, JMB, CHT, MM, and AV acquired data. MH, SPL, CHT, and AV analyzed data. SPL, CHT, and AV wrote the manuscript. All authors reviewed and revised the manuscript.

## Acknowledgments

The authors thank the Mass General Brigham research team for their support with participant recruitment, study coordination, data collection, and research operations.

