## Supplementals for "Reprogramming of Iron and Oxygen Metabolism Across the Spectrum of Primary Aldosteronism"

### Supplemental Figure Legends:

**Supplemental Figure 1: The Proteomic Signature of Primary Aldosteronism**

Volcano plot showing 903 proteins differentially abundant between normotensive individuals (blue) and those with Overt PA (red).

**Supplemental Figure 2: The Proteomic Signature of Primary Aldosteronism After Adjustment for Age, Sex, Body Mass Index, Systolic Blood Pressure, Serum Potassium, and Glomerular Filtration Rate**

Volcano plot showing 526 proteins differentially abundant between normotensive individuals (blue) and those with Overt PA (red).

**Supplemental Figure 3: Reactome Pathway Enrichment and Differential Abundance of Iron and Heme-Related Proteins in Primary Aldosteronism After Adjustment for Age, Sex, Body Mass Index, Systolic Blood Pressure, Serum Potassium, and Glomerular Filtration Rate**

A) Reactome pathway enrichment of differentially abundant proteins filtered for iron and heme-related terms, yielding 11 pathways ($p<0.1$). Dot size reflects the number of proteins per pathway and color indicates p-value.

B) Volcano plot of proteins belonging to these 11 iron and heme-related pathways. Blue dots indicate proteins higher in PA; red dots indicate proteins higher in normotensive (NT) controls.

**Supplemental Figure 4: Protein-by-Pathway Heatmap of Iron and Heme-Related Proteins in Primary Aldosteronism After Adjustment for Age, Sex, Body Mass Index, Systolic Blood Pressure, Serum Potassium, and Glomerular Filtration Rate.**

Each row represents a differentially abundant protein and each column a Reactome iron or heme-related pathway. Cell color reflects the log fold change (logFC) of each protein in PA versus normotensive controls (red = higher in PA, blue = lower in PA). Black dots indicate statistically significant associations ($p<0.05$). Only proteins belonging to at least one of the 11 enriched pathways are shown.

**Supplemental Figure 5: Iron and Heme-Related Plasma Proteins Across the Primary Aldosteronism Continuum After Adjustment for Age, Sex, Body Mass Index, Systolic Blood Pressure, Serum Potassium, and Glomerular Filtration Rate**

A) Percent change in plasma protein levels relative to subclinical PA tertile 1 (T1) across increasing aldosterone exposure. Blue and red lines indicate proteins with upward and downward trends, respectively. Gray bars represent mean plasma aldosterone (right y-axis).

B) Functional annotation of the trending proteins, highlighting enrichment for heme biosynthesis, oxygen transport, and mitochondrial electron transport functions.

**Supplemental Figure 6. Association Between Plasma Aldosterone Concentration and Hemoglobin in an Independent Hypertensive Cohort.**

Hemoglobin levels across quartiles of plasma aldosterone concentration in an independent Taiwanese cohort of 5,713 patients with hypertension. Plasma aldosterone concentration increased from Q1 to Q4, with median values of 8.1, 15.9, 26.3, and 44.9 ng/dL, respectively. Hemoglobin levels increased progressively across aldosterone quartiles, from 12.3 ± 2.4 g/dL in Q1 to 13.3 ± 2.0 g/dL in Q4. The trend remained significant after multivariable adjustment with age, eGFR, sex body mass index, number of hypertensive medications (adjusted P for trend <0.001).

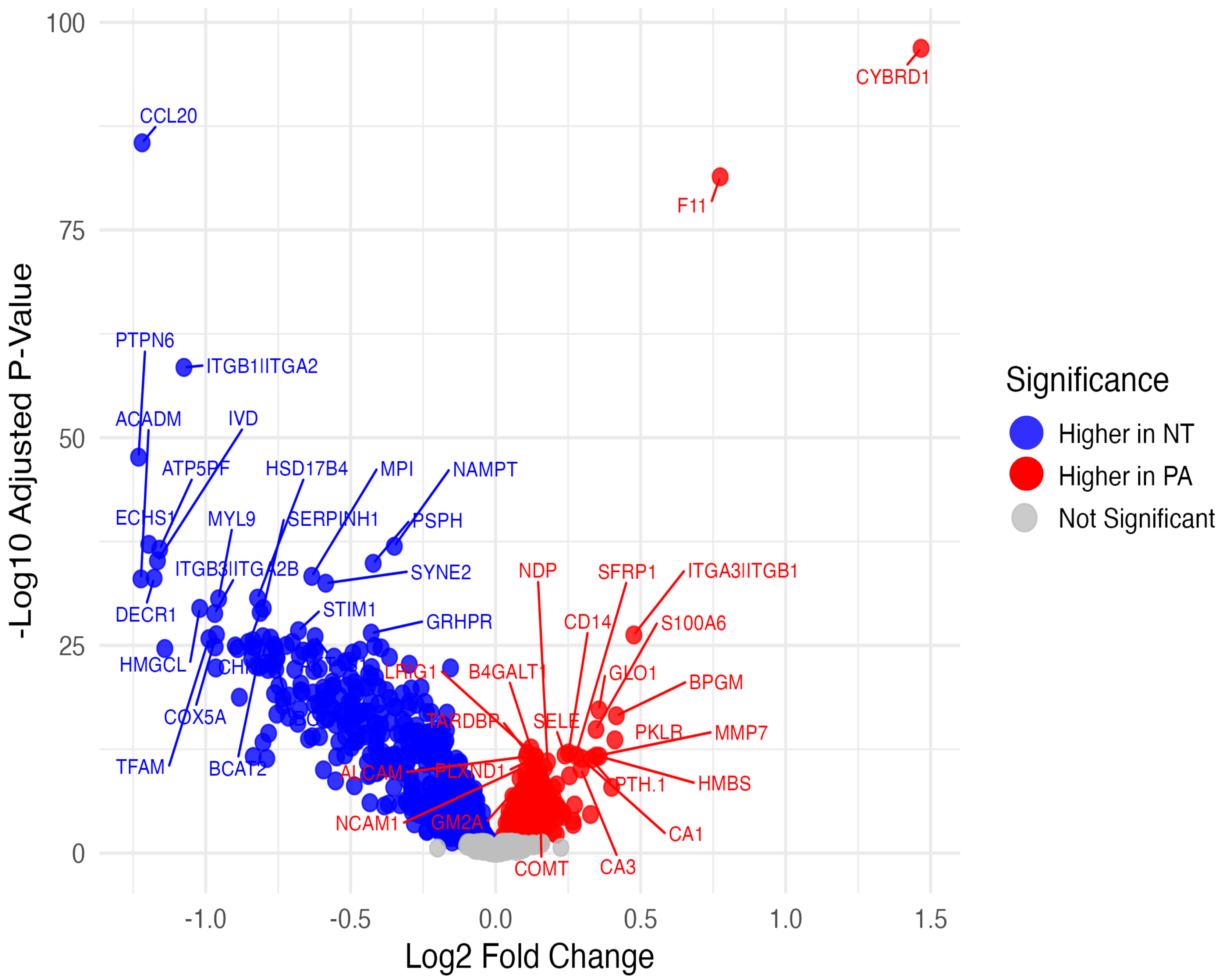

-Log10 Adjusted P-Value
Log2 Fold Change
100
75
50
25
0
-1.0
-0.5
0.0
0.5
1.0
1.5
Significance
Higher in NT
Higher in PA
Not Significant
CYBRD1
CCL20
F11
PTPN6
ITGB1|ITGA2
ACADM
IVD
ATP5PF
HSD17B4
MPI
NAMPT
ECHS1
MYL9
SERPINH1
PSPH
ITGB3|ITGA2B
SYNE2
NDP
SFRP1
ITGA3|ITGB1
DECR1
STIM1
GRHPR
CD14
S100A6
HMGCL
LRIG1
B4GALT1
GLO1
BPGM
COX5A
TARDBP
SELE
PKLR
MMP7
TFAM
BCAT2
ALCAM
PLXND1
PTH.1
HMBS
NCAM1
GM2A
CA1
COMT
CA3

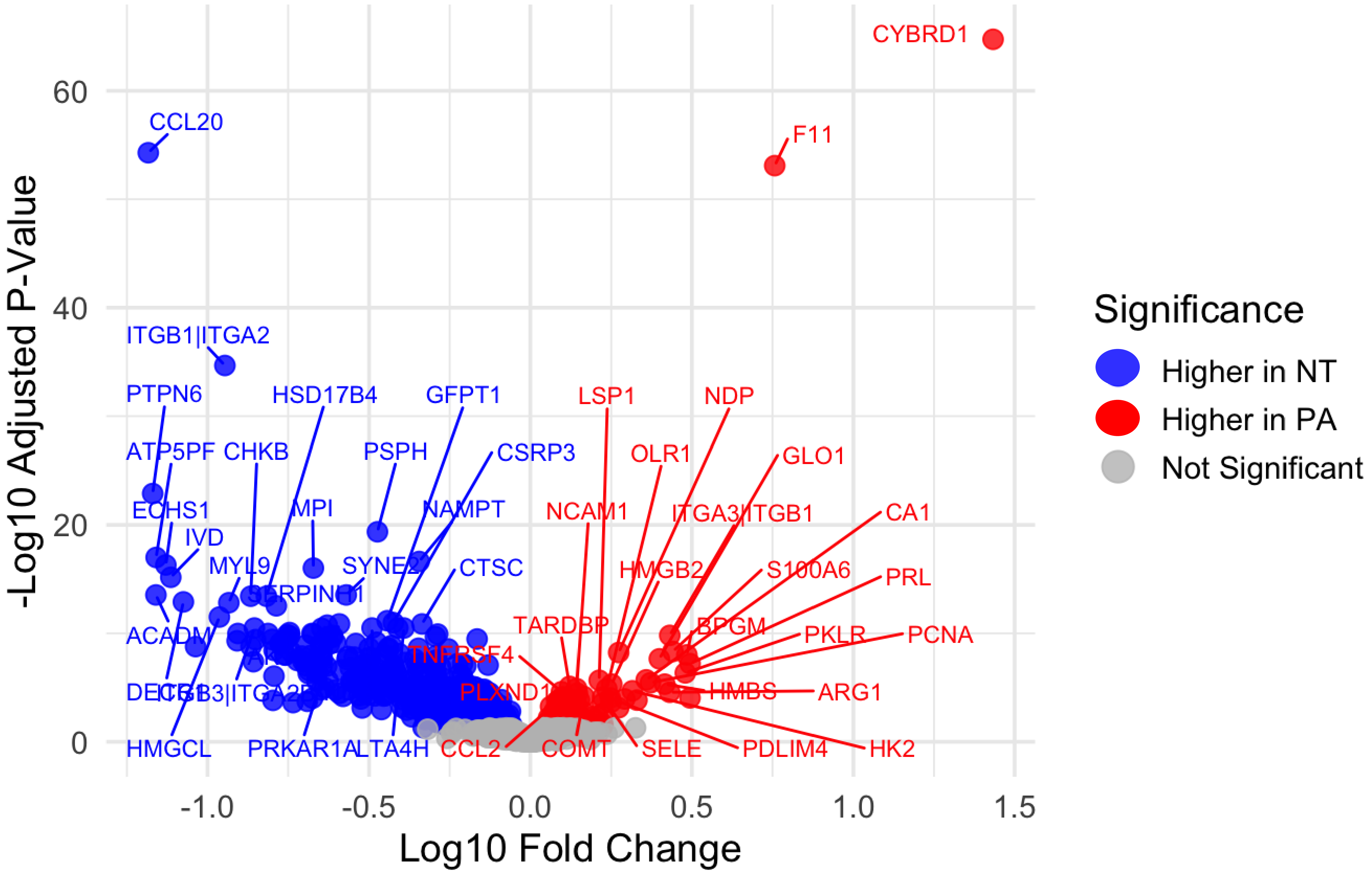

-Log10 Adjusted P-Value
Log10 Fold Change
Significance
Higher in NT
Higher in PA
Not Significant
CYBRD1
F11
CCL20
ITGB1|ITGA2
PTPN6
HSD17B4
GFPT1
LSP1
NDP
ATP5PF
CHKB
PSPH
CSRP3
OLR1
GLO1
ECHS1
MPI
NAMPT
NCAM1
ITGA3|ITGB1
CA1
IVD
MYL9
SYNE2
CTSC
HMGB2
S100A6
PRL
SERPINH1
TARDBP
BPGM
PKLR
PCNA
ACADM
TNFRSF4
PLXND1
HMBS
ARG1
HMGCL
PRKAR1A
LTA4H
CCL2
COMT
SELE
PDLIM4
HK2

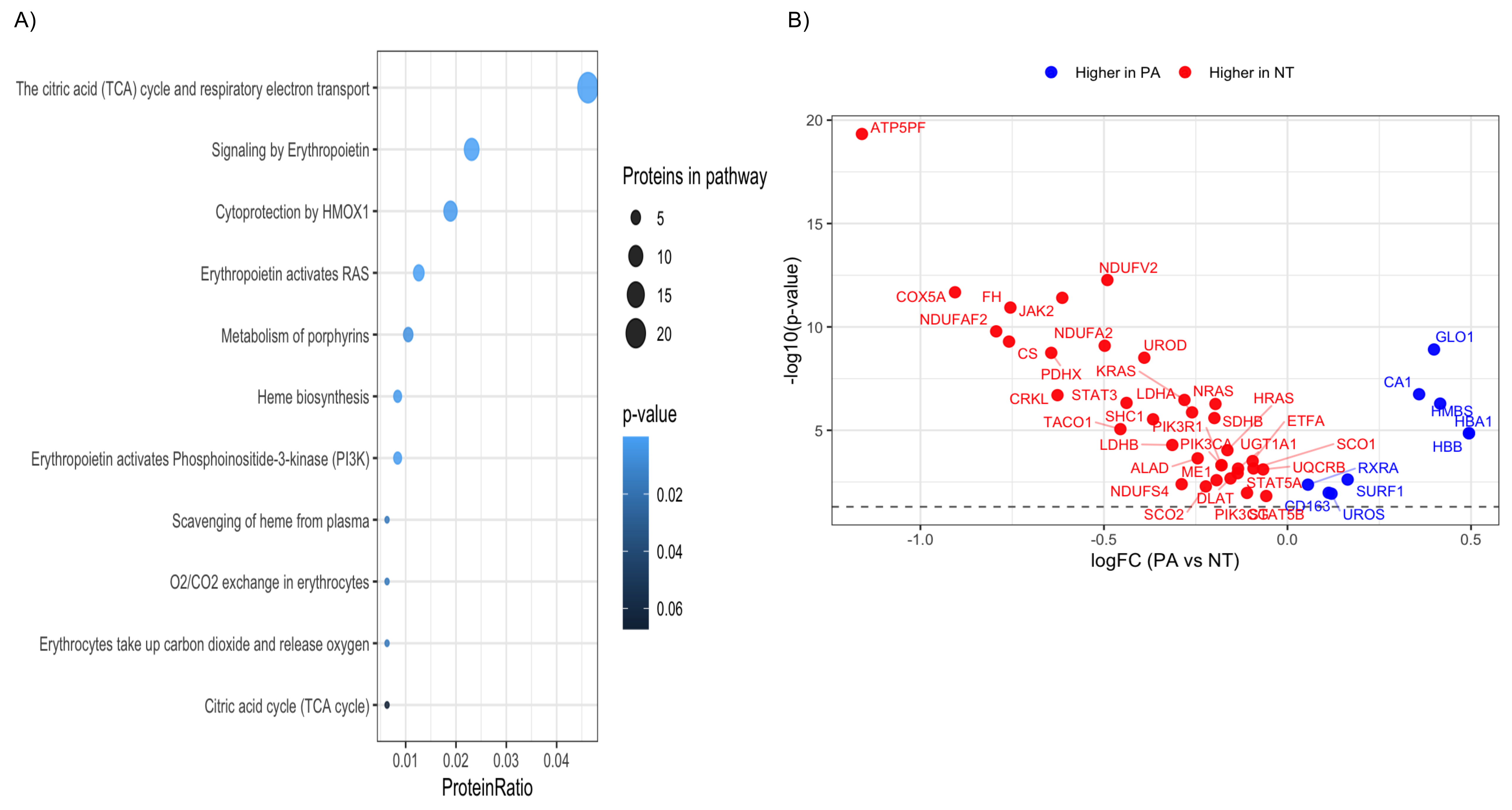
A)
The citric acid (TCA) cycle and respiratory electron transport
Signaling by Erythropoietin
Cytoprotection by HMOX1
Erythropoietin activates RAS
Metabolism of porphyrins
Heme biosynthesis
Erythropoietin activates Phosphoinositide-3-kinase (PI3K)
Scavenging of heme from plasma
O2/CO2 exchange in erythrocytes
Erythrocytes take up carbon dioxide and release oxygen
Citric acid cycle (TCA cycle)
0.01
0.02
0.03
0.04
ProteinRatio
Proteins in pathway
5
10
15
20
p-value
0.02
0.04
0.06
B)
Higher in PA
Higher in NT
-log10(p-value)
20
15
10
5
logFC (PA vs NT)
-1.0
-0.5
0.0
0.5
ATP5PF
NDUFV2
COX5A
FH
JAK2
NDUFAF2
NDUFA2
UROD
CS
PDHX
KRAS
CRKL
STAT3
LDHA
NRAS
HRAS
SHC1
SDHB
ETFA
TACO1
PIK3R1
LDHB
PIK3CA
UGT1A1
SCO1
ALAD
ME1
UQCRB
RXRA
NDUFS4
STAT5A
SURF1
DLAT
CD163
SCO2
PIK3CG
STAT5B
UROS
GLO1
CA1
HMBS
HBA1
HBB

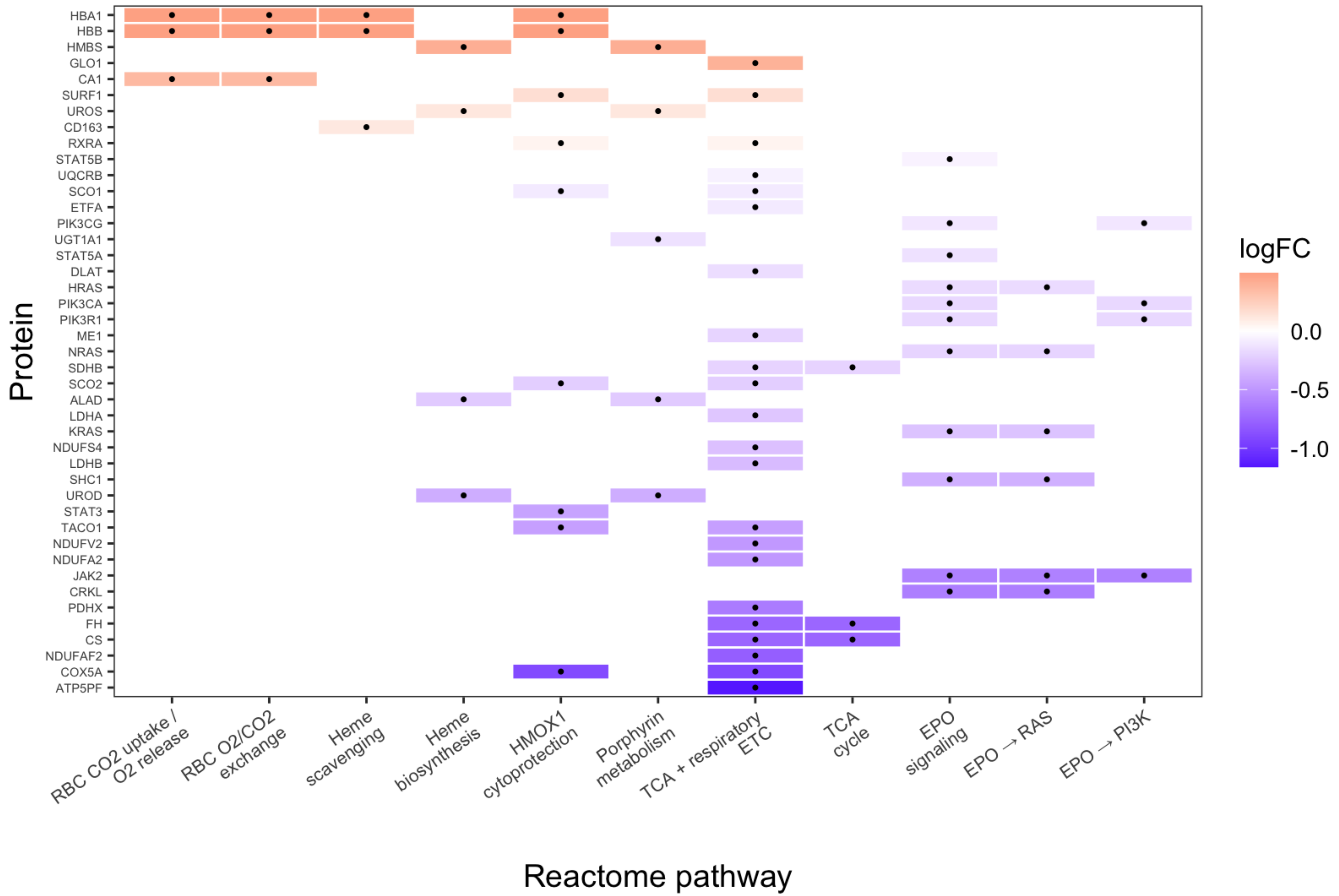

Protein
HBA1
HBB
HMBS
GLO1
CA1
SURF1
UROS
CD163
RXRA
STAT5B
UQCRB
SCO1
ETFA
PIK3CG
UGT1A1
STAT5A
DLAT
HRAS
PIK3CA
PIK3R1
ME1
NRAS
SDHB
SCO2
ALAD
LDHA
KRAS
NDUFS4
LDHB
SHC1
UROD
STAT3
TACO1
NDUFV2
NDUFA2
JAK2
CRKL
PDHX
FH
CS
NDUFAF2
COX5A
ATP5PF
RBC CO2 uptake / O2 release
RBC O2/CO2 exchange
Heme scavenging
Heme biosynthesis
HMOX1 cytoprotection
Porphyrin metabolism
TCA + respiratory ETC
TCA cycle
EPO signaling
EPO → RAS
EPO → PI3K
Reactome pathway
logFC
0.0
-0.5
-1.0

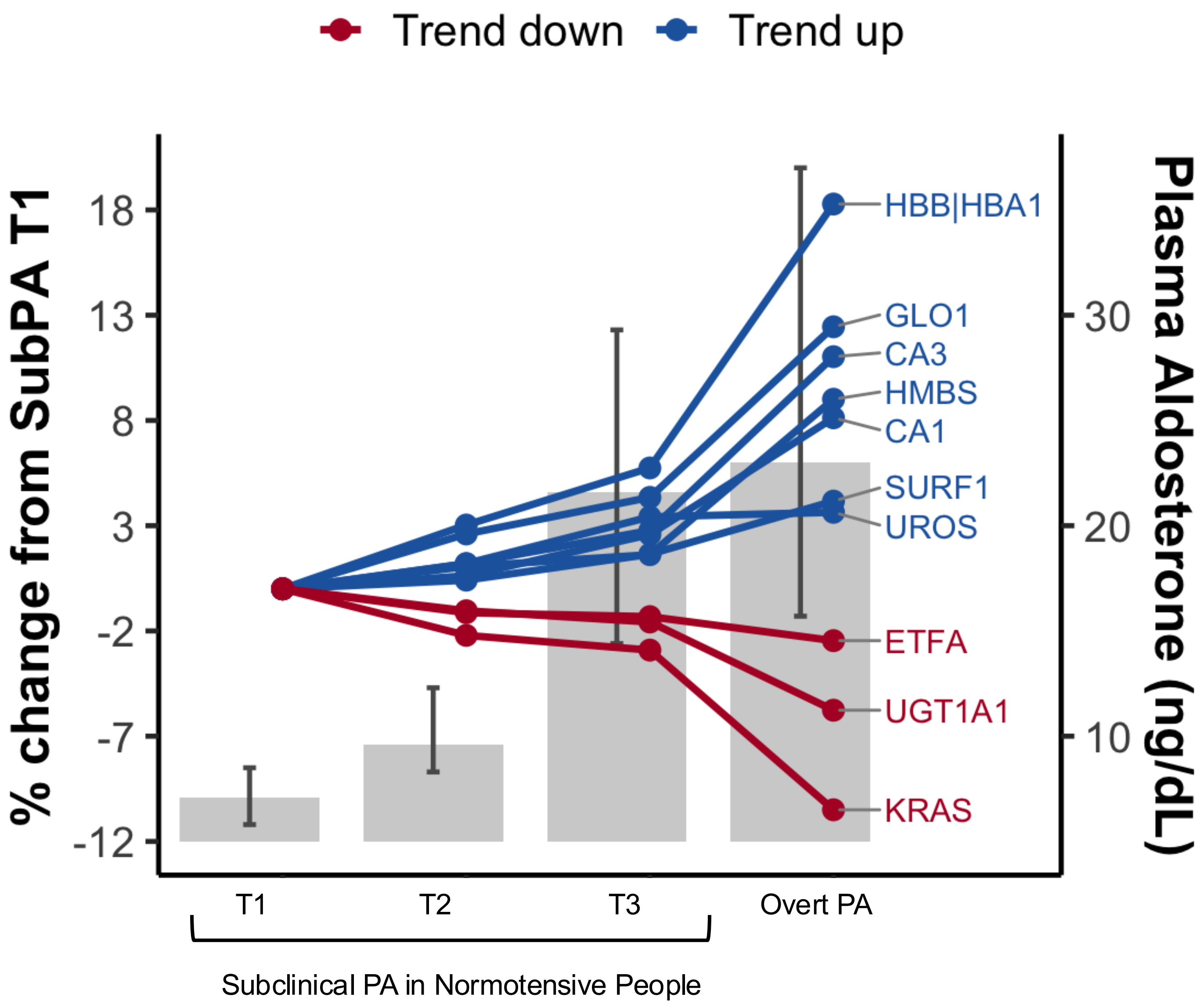


| Protein | Direction Across PA Continuum | Function |
|---|---|---|
| **HBA1 / HBB** | Upward | Hemoglobin β and α chains: oxygen transport in red blood cells |
| **GLO1** | Upward | Methylglyoxal detoxification enzyme. |
| **CA3** | Upward | Involved in $CO_2$ handling and red cell metabolism |
| **HMBS** | Upward | Enzyme in heme biosynthesis (porphyrin pathway) |
| **CA1** | Upward | Mediates $CO_2$/$O_2$ exchange in erythrocytes |
| **SURF1** | Upward | Cytochrome c oxidase (Complex IV) assembly factor |
| **UROS** | Upward | Heme biosynthesis (uroporphyrinogen synthesis) |
| **ETFA** | Downward | Electron transfer to mitochondrial respiratory chain |
| **UGT1A1** | Downward | Bilirubin conjugation and hepatic heme catabolite clearance |
| **KRAS** | Downward | RAS GTPase |

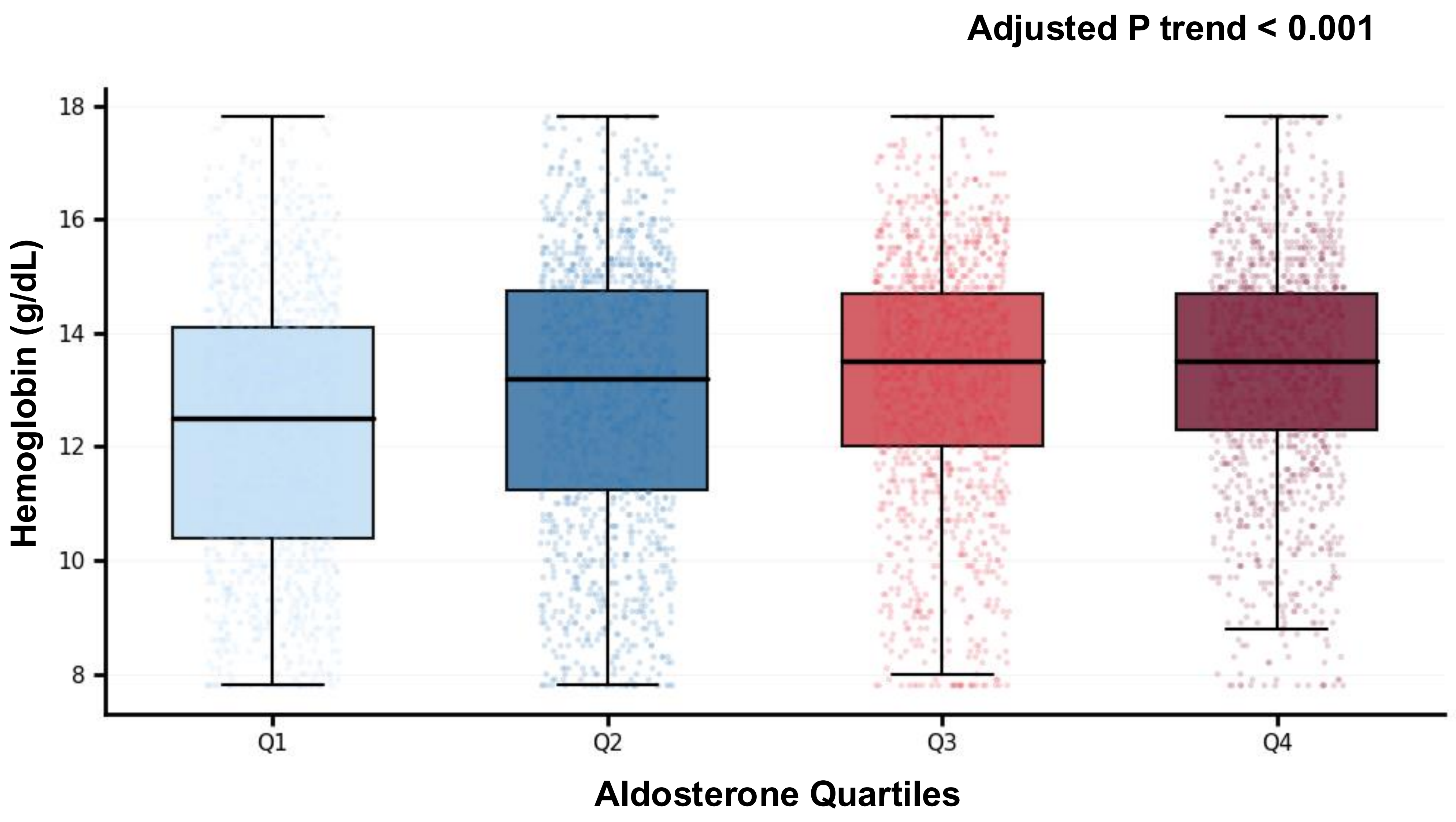
Adjusted P trend < 0.001
Hemoglobin (g/dL)
18
16
14
12
10
8
Q1
Q2
Q3
Q4
Aldosterone Quartiles

| SeqId | Target | GeneSymbol |
|---|---|---|
| 10001-7 | c-Raf | RAF1 |
| 10011-65 | OCRL | OCRL |
| 10014-31 | SLUG | SNAI2 |
| 10022-207 | POLH | POLH |
| 10023-32 | VDR | VDR |
| 10024-44 | HOGA1 | HOGA1 |
| 10041-3 | HNF4A | HNF4A |
| 10043-31 | BRD4 | BRD4 |
| 10048-7 | PEBB | CBFB |
| 10053-5 | ILK1 | ILK |
| 10063-10 | FANCL | FANCL |
| 10078-5 | BAG3 | BAG3 |
| 10085-25 | STAR | STAR |
| 10086-39 | CBS | CBS |
| 10088-37 | APT | APRT |
| 10346-5 | STAT3 | STAT3 |
| 10351-51 | IRF1 | IRF1 |
| 10356-21 | c-Jun | JUN |
| 10362-35 | c-Myc | MYC |
| 10363-13 | SMAD3 | SMAD3 |
| 10364-6 | SMAD2 | SMAD2 |
| 10365-132 | IL-23 | IL12B\|IL23A |
| 10366-11 | PDGFRA | PDGFRA |
| 10367-62 | IL-12 | IL12A\|IL12B |
| 10391-1 | ANGL3 | ANGPTL3 |
| 10396-6 | Mcl-1 | MCL1 |
| 10426-21 | GOSR2 | GOSR2 |
| 10435-2 | ASPG | AGA |
| 10480-33 | CD59 | CD59 |
| 10521-10 | MXRA8:ECD | MXRA8 |
| 10531-18 | RASN | NRAS |
| 10534-40 | PARP:BRCT domain | PARP1 |
| 10550-37 | BMP RIB | BMPR1B |
| 10551-7 | LAT | LAT |
| 10552-88 | NKG2F | KLRC4 |
| 10554-23 | BGAL | GLB1 |
| 10574-10 | b2-Microglobulin | B2M |
| 10584-7 | NDUS4 | NDUFS4 |
| 10618-190 | megalin | LRP2 |
| 10623-19 | MUC1:region 1 | MUC1 |
| 10627-87 | APLP2 | APLP2 |
| 10638-1 | TINF2 | TINF2 |
| 10666-7 | GNPTG | GNPTG |
| 10667-78 | CA185 | C1orf185 |
| 10710-23 | ZPBP2 | ZPBP2 |
| 10714-7 | ACE | ACE |

| | | |
|---|---|---|
| 10761-5 | TMED2 | TMED2 |
| 10800-15 | Collagen-binding prote | SERPINH1 |
| 10818-36 | ASM | SMPD1 |
| 10889-2 | MRAP2 | MRAP2 |
| 10892-8 | OSMR | OSMR |
| 10945-11 | Syntaxin-6 | STX6 |
| 10955-4 | CLC10 | CLEC10A |
| 10956-82 | GCSH | GCSH |
| 10966-1 | a2-HS-Glycoprotein | AHSG |
| 10976-44 | MUC1:region 3 | MUC1 |
| 10980-11 | ACES | ACHE |
| 11067-13 | Osteocalcin | BGLAP |
| 11081-1 | GPDA | GPD1 |
| 11101-18 | TLR4 | TLR4 |
| 11103-24 | HSP 27 | HSPB1 |
| 11104-13 | YKL-40 | CHI3L1 |
| 11105-171 | Alpha enolase | ENO1 |
| 11138-16 | RUNX3 | RUNX3 |
| 11140-56 | CO1A1:C-term propep | COL1A1 |
| 11171-25 | filamin A:CH2 | FLNA |
| 11185-145 | GCH1 | GCH1 |
| 11193-27 | HNF1A | HNF1A |
| 11201-19 | TRUA | PUS1 |
| 11202-70 | TBX5 | TBX5 |
| 11203-97 | KPYR | PKLR |
| 11208-15 | NAGPA | NAGPA |
| 11211-7 | TBCE | TBCE |
| 11218-84 | TPMT | TPMT |
| 11231-12 | ADXL | FDX2 |
| 11237-49 | PCOC1 | PCOLCE |
| 11239-49 | TMPS6 | TMPRSS6 |
| 11241-8 | ARLY | ASL |
| 11242-33 | TGM1 | TGM1 |
| 11245-43 | filamin A:CH1 | FLNA |
| 11247-20 | NAGS | NAGS |
| 11248-43 | HEM4 | UROS |
| 11257-1 | DHPR | QDPR |
| 11264-33 | XDH | XDH |
| 11266-8 | SELPL:ECD | SELPLG |
| 11270-17 | CHC10 | CHCHD10 |
| 11287-14 | Cytochrome b5 | CYB5A |
| 11297-54 | NOTC2 | NOTCH2 |
| 11300-32 | SORT | SORT1 |
| 11303-7 | SAMH1 | SAMHD1 |
| 11311-79 | RAG-1 | RAG1 |
| 11312-40 | PMS2 | PMS2 |
| 11313-100 | PHS | PCBD1 |

| | | |
|---|---|---|
| 11319-106 | MRE11 | MRE11 |
| 11336-9 | FANCF | FANCF |
| 11338-49 | BLK | BLK |
| 11347-9 | Transaldolase | TALDO1 |
| 11351-233 | NHEJ1 | NHEJ1 |
| 11352-42 | TITIN | TTN |
| 11356-19 | DGC14 | ESS2 |
| 11364-18 | MFN1 | MFN1 |
| 11368-32 | KAD2 | AK2 |
| 11370-20 | INP5E | INPP5E |
| 11381-56 | PRPS1 | PRPS1 |
| 11383-41 | Keratin 7 | KRT7 |
| 11391-69 | MVK | MVK |
| 11396-39 | DNAI1 | DNAI1 |
| 11406-82 | ACAD8 | ACAD8 |
| 11424-4 | FAAA | FAH |
| 11432-11 | PQBP1 | PQBP1 |
| 11436-6 | AINX | INA |
| 11440-58 | SOCS-3 | SOCS3 |
| 11441-11 | PYGL | PYGL |
| 11448-34 | GALK1 | GALK1 |
| 11450-110 | Protein disulfide-isom | P4HB |
| 11457-53 | GALE | GALE |
| 11516-7 | FABPL | FABP1 |
| 11530-37 | HEM3 | HMBS |
| 11537-12 | TFR2 | TFR2 |
| 11538-216 | DCMC | MLYCD |
| 11540-37 | FOXO3A | FOXO3 |
| 11616-9 | HSF1 | HSF1 |
| 11617-1 | LFA-1 alpha-L chain | ITGAL |
| 11696-7 | RABP2 | CRABP2 |
| 11699-16 | TP4A2 | PTP4A2 |
| 11709-29 | CPT1B | CPT1B |
| 11814-29 | Met | MET |
| 11816-84 | JAK2 | JAK2 |
| 11825-27 | PRGC1 | PPARGC1A |
| 11830-48 | SHP-2 | PTPN11 |
| 11833-83 | FRDA | FXN |
| 11838-130 | FA38A | PIEZO1 |
| 11949-25 | EGF:CD | EGF |
| 12008-3 | CD7 | CD7 |
| 12014-19 | PTPS | PTS |
| 12016-60 | CBL | CBL |
| 12020-39 | PMGE | BPGM |
| 12022-12 | SMAD4 | SMAD4 |
| 12030-82 | Desmin | DES |
| 12034-28 | CAP 1 | CAP1 |

| | | |
|---|---|---|
| 12046-51 | TADBP | TARDBP |
| 12077-32 | Myostatin | MSTN |
| 12333-87 | RPIA | RPIA |
| 12341-8 | DUS6 | DUSP6 |
| 12347-29 | CCM2 | CCM2 |
| 12348-46 | SYSM | SARS2 |
| 12351-25 | STAT1 | STAT1 |
| 12361-102 | RRAS2 | RRAS2 |
| 12367-52 | MACOI | MACO1 |
| 12378-71 | TPSN | TAPBP |
| 12381-26 | CBR1 | CBR1 |
| 12382-2 | DDX58 | DDX58 |
| 12387-7 | PDLI4 | PDLIM4 |
| 12395-86 | SYDM | DARS2 |
| 12396-19 | HIBCH | HIBCH |
| 12398-15 | PAX4 | PAX4 |
| 12400-25 | UBE2T | UBE2T |
| 12409-90 | RAB7B | RAB7B |
| 12420-10 | GPD1L | GPD1L |
| 12427-8 | MPIP2 | CDC25B |
| 12444-39 | NR5A2 | NR5A2 |
| 12446-49 | GST A1-1 | GSTA1 |
| 12456-5 | F263 | PFKFB3 |
| 12459-13 | PKHA1 | PLEKHA1 |
| 12466-7 | ROA1 | HNRNPA1 |
| 12469-19 | MARE1 | MAPRE1 |
| 12488-9 | MCTS1 | MCTS1 |
| 12501-10 | TBCA | TBCA |
| 12504-26 | LMOD1 | LMOD1 |
| 12507-16 | IP3KC | ITPKC |
| 12509-115 | COMD1 | COMMD1 |
| 12510-3 | STAP1 | STAP1 |
| 12521-3 | CDN2C | CDKN2C |
| 12527-50 | THA | THRA |
| 12529-32 | FKBP6 | FKBP6 |
| 12538-19 | RGS7 | RGS7 |
| 12574-36 | Endothelin 2 | EDN2 |
| 12580-7 | PSB5 | PSMB5 |
| 12585-39 | ERCC1 | ERCC1 |
| 12604-16 | SCMH1 | SCMH1 |
| 12612-37 | PSB1 | PSMB1 |
| 12620-3 | Septin-11 | SEPTIN11 |
| 12650-43 | GNAI3 | GNAI3 |
| 12651-21 | PDK2 | PDK2 |
| 12687-2 | DECR | DECR1 |
| 12700-9 | ACLY | ACLY |
| 12705-9 | HERC1 | HERC1 |

| | | |
|---|---|---|
| 12712-9 | HM20A | HMG20A |
| 12720-71 | UBQL4 | UBQLN4 |
| 12724-81 | CIRBP | CIRBP |
| 12726-3 | RFXAP | RFXAP |
| 12738-43 | NISCH | NISCH |
| 12740-55 | FEV | FEV |
| 12756-3 | BACH2 | BACH2 |
| 12825-18 | DP13A | APPL1 |
| 12869-68 | DDX6 | DDX6 |
| 12873-11 | LEUK | SPN |
| 12895-28 | DGKB | DGKB |
| 12904-180 | cubilin | CUBN |
| 12931-16 | MCR | NR3C2 |
| 12954-71 | PPARa | PPARA |
| 12957-62 | UCR6 | UQCRB |
| 12960-9 | HXK4 | GCK |
| 12988-49 | EWS | EWSR1 |
| 13027-20 | CBX7 | CBX7 |
| 13032-1 | BECN1 | BECN1 |
| 13053-6 | S6A14 | SLC6A14 |
| 13067-5 | KGP1B | PRKG1 |
| 13085-18 | GLP1R:CD | GLP1R |
| 13088-397 | BTC | BTC |
| 13089-6 | HIF-1a | HIF1A |
| 13090-17 | S100A6 | S100A6 |
| 13098-93 | VEGF-D | VEGFD |
| 13105-7 | SNP25 | SNAP25 |
| 13112-179 | FSTL1 | FSTL1 |
| 13113-7 | Osteopontin | SPP1 |
| 13119-26 | protein Z inhibitor | SERPINA10 |
| 13126-52 | DSC2 | DSC2 |
| 13129-40 | LDLR | LDLR |
| 13130-150 | HXK2 | HK2 |
| 13131-5 | HXK1 | HK1 |
| 13236-25 | WNT3A | WNT3A |
| 13381-49 | B4GT1 | B4GALT1 |
| 13384-110 | FUMH | FH |
| 13388-57 | NEC1 | PCSK1 |
| 13392-13 | AT1B1 | ATP1B1 |
| 13405-61 | ISK2 | SPINK2 |
| 13451-2 | FNDC4 | FNDC4 |
| 13465-5 | CCP1 | RCAN1 |
| 13470-43 | PTH1R | PTH1R |
| 13484-69 | CO1A1:N-term propep | COL1A1 |
| 13486-9 | PKD2:CD 1 | PKD2 |
| 13492-44 | HSPA9B | HSPA9 |
| 13499-30 | Coagulation Factor VIII | F8 |

| | | |
|---|---|---|
| 13510-7 | AT2A3 | ATP2A3 |
| 13556-28 | 5HT2A | HTR2A |
| 13576-15 | Glutathione S-transfer | GSTP1 |
| 13604-27 | NEUL1 | NEURL1 |
| 13609-11 | GTF2I | GTF2I |
| 13614-6 | CREB-binding protein | CREBBP |
| 13628-58 | GRB14 | GRB14 |
| 13634-209 | PIR | PIR |
| 13654-1 | ROCK2 | ROCK2 |
| 13655-34 | Nucleolin | NCL |
| 13657-2 | PNKP | PNKP |
| 13665-35 | PP2A, subunit B | PPP2R3A |
| 13669-6 | FGFR-3:ECD | FGFR3 |
| 13676-46 | Inhibin bB chain | INHBB |
| 13678-169 | Factor D | CFD |
| 13698-28 | K319L | KIAA0319L |
| 13700-10 | annexin II | ANXA2 |
| 13704-5 | HMCS2 | HMGCS2 |
| 13717-15 | FCN2 | FCN2 |
| 13726-4 | B7 | CD80 |
| 13730-18 | CATC | CTSC |
| 13732-79 | Cardiotrophin-1 | CTF1 |
| 13733-5 | IL-12 p40 | IL12B |
| 13741-36 | IGFBP-1 | IGFBP1 |
| 13745-10 | PKD2:CD 4 | PKD2 |
| 13941-82 | SYFM | FARS2 |
| 13967-14 | Thioredoxin reductase | TXNRD1 |
| 13977-28 | BARD1 | BARD1 |
| 13978-122 | Tbx3 | TBX3 |
| 13990-1 | pyruvate carboxylase | PC |
| 14005-2 | CHD7 | CHD7 |
| 14006-36 | GNMT | GNMT |
| 14025-18 | 4-1BB | TNFRSF9 |
| 14030-21 | OX40 Ligand | TNFSF4 |
| 14031-18 | FGF7 | FGF7 |
| 14035-13 | TIAM1 | TIAM1 |
| 14047-78 | BDNF | BDNF |
| 14051-54 | FOXC2 | FOXC2 |
| 14052-26 | UN13A | UNC13A |
| 14060-67 | NCF4 | NCF4 |
| 14061-48 | sRANKL | TNFSF11 |
| 14066-49 | MAGI2 | MAGI2 |
| 14067-6 | PKP2 | PKP2 |
| 14069-61 | Carbonic Anhydrase IV | CA4 |
| 14094-29 | HB-EGF | HBEGF |
| 14105-5 | Kallistatin | SERPINA4 |
| 14110-200 | ON | SPARC |

| | | |
|---|---|---|
| 14131-37 | EFNB2:ECD | EFNB2 |
| 14136-234 | C1QR1 | CD93 |
| 14158-17 | Annexin V | ANXA5 |
| 14284-23 | E2AK4 | EIF2AK4 |
| 14645-253 | GSTA4 | GSTA4 |
| 14756-29 | sICAM-2 | ICAM2 |
| 15295-81 | IGF-II:Mature | IGF2 |
| 15299-102 | MESD2 | MESD |
| 15303-63 | S100A5 | S100A5 |
| 15324-58 | Ferritin light chain | FTL |
| 15339-32 | COF2 | CFL2 |
| 15343-337 | Kininogen, HMW, Two | KNG1 |
| 15346-31 | IFN-g | IFNG |
| 15361-37 | ANKR1 | ANKRD1 |
| 15363-32 | Apo A-V | APOA5 |
| 15364-101 | Apo C-I | APOC1 |
| 15383-200 | Endothelin 3 | EDN3 |
| 15384-15 | KLOTHO | KL |
| 15385-116 | FABP2 | FABP2 |
| 15386-7 | FABPA | FABP4 |
| 15388-24 | FcRIIIa | FCGR3A |
| 15390-3 | Galanin | GAL |
| 15391-114 | GAS-6 | GAS6 |
| 15395-15 | GST M1-1 | GSTM1 |
| 15398-2 | HERV1 | GFER |
| 15413-3 | LCAT | LCAT |
| 15414-316 | LDHA | LDHA |
| 15431-31 | OTCase | OTC |
| 15433-4 | p60-Src | SRC |
| 15434-5 | PNOC | PNOC |
| 15435-4 | PNP | PNP |
| 15440-57 | NEC2 | PCSK2 |
| 15441-6 | SAP3 | GM2A |
| 15447-45 | Sorbitol dehydrogenas | SORD |
| 15448-47 | SQSTM | SQSTM1 |
| 15449-33 | TIM-4 | TIMD4 |
| 15452-5 | 5'-Nucleotidase | NT5E |
| 15453-3 | a1-Microglobulin | AMBP |
| 15457-14 | Aminopeptidase N | ANPEP |
| 15460-9 | CD26 | DPP4 |
| 15467-10 | CTHR1 | CTHRC1 |
| 15470-11 | Hexosaminidase B | HEXB |
| 15475-4 | PLTP | PLTP |
| 15476-6 | REG3G | REG3G |
| 15482-12 | A2ML1 | A2ML1 |
| 15486-126 | ABP1 | AOC1 |
| 15503-20 | Lefty-A | LEFTY2 |

| | | |
|---|---|---|
| 15509-2 | NAG | NAGLU |
| 15513-108 | Prostasin | PRSS8 |
| 15514-26 | Pseudocholinesterase | BCHE |
| 15515-2 | SAA | SAA1 |
| 15523-9 | HEM2 | ALAD |
| 15524-30 | PGAM2 | PGAM2 |
| 15526-33 | GSHB | GSS |
| 15527-90 | DLDH | DLD |
| 15530-33 | EphB4 | EPHB4 |
| 15535-3 | Marapsin | PRSS27 |
| 15540-6 | Vimentin | VIM |
| 15545-13 | Calcineurin B a | PPP3R1 |
| 15559-5 | ANTR2 | ANTXR2 |
| 15562-24 | BGLR | GUSB |
| 15567-2 | CD3-zeta | CD247 |
| 15570-99 | Complement receptor | CR2 |
| 15585-304 | fibulin 5 | FBLN5 |
| 15588-17 | galactosidase, alpha | GLA |
| 15589-1 | Gc-Globulin, Mixed Ty | GC |
| 15591-28 | Glutathione peroxidas | GPX1 |
| 15594-47 | HTRA1 | HTRA1 |
| 15603-20 | Integrin alpha-2 | ITGA2 |
| 15604-18 | JNK2 | MAPK9 |
| 15606-19 | Keratin 19 | KRT19 |
| 15608-5 | KS6B1 | RPS6KB1 |
| 15626-223 | Perlecan | HSPG2 |
| 15633-6 | RBP | RBP4 |
| 15635-4 | SMOC2 | SMOC2 |
| 15666-21 | BMP-2 | BMP2 |
| 15667-39 | BMP-4 | BMP4 |
| 15669-7 | BRAF1 | BRAF |
| 15692-300 | NODAL | NODAL |
| 16035-8 | VEGF sR3 | FLT4 |
| 16043-30 | SHC1:PID | SHC1 |
| 16057-6 | IGF-II receptor | IGF2R |
| 16288-17 | EPHA4 | EPHA4 |
| 16297-14 | ROBO4 | ROBO4 |
| 16318-12 | ALK-1 | ACVRL1 |
| 16320-139 | CD320 | CD320 |
| 16323-8 | NRX3A | NRXN3 |
| 16536-3 | HMGA1 | HMGA1 |
| 16551-14 | STA5B | STAT5B |
| 16588-10 | BiP | HSPA5 |
| 16593-3 | FADD | FADD |
| 16596-25 | GLRX3 | GLRX3 |
| 16597-11 | GLRX5 | GLRX5 |
| 16612-28 | SDC3 | SDC3 |

| | | |
|---|---|---|
| 16616-137 | ENOB | ENO3 |
| 16618-7 | CD69 | CD69 |
| 16754-40 | CSKP | CASK |
| 16768-3 | PLAK | JUP |
| 16803-4 | CALB2 | CALB2 |
| 16809-1 | NDKM | NME4 |
| 16814-13 | SLIM 1 | FHL1 |
| 16828-8 | Collagen a1(VI) | COL6A1 |
| 16850-5 | RGS5 | RGS5 |
| 16851-50 | SCO2 | SCO2 |
| 16865-62 | RBP56 | TAF15 |
| 16890-37 | ATL1 | ADAMTSL1 |
| 16914-104 | sCD14 | CD14 |
| 16915-153 | SEM4A | SEMA4A |
| 16916-19 | SLIK6 | SLITRK6 |
| 16926-44 | Alkaline phosphatase, | ALPL |
| 16927-9 | Coagulation factor XIII | F13B\|F13A1 |
| 17145-1 | S100A8/S100A9 | S100A9\|S100A8 |
| 17150-8 | HSP 10 | HSPE1 |
| 17161-1 | OST48 | DDOST |
| 17205-21 | RAB5A | RAB5A |
| 17333-20 | ACADM | ACADM |
| 17341-89 | THIC | ACAT2 |
| 17350-13 | CHM2B | CHMP2B |
| 17362-5 | VHL | VHL |
| 17377-1 | Aldose reductase-like ( | AKR1C3 |
| 17384-110 | K6PF | PFKM |
| 17393-13 | SCAD | ACADS |
| 17398-55 | HO-1 | HMOX1 |
| 17403-14 | acyl-Coenzyme A dehy | ACADSB |
| 17411-55 | UFD1 | UFD1 |
| 17435-43 | ETFA | ETFA |
| 17447-52 | SFRP4 | SFRP4 |
| 17451-13 | BIM | BCL2L11 |
| 17453-34 | Ceruloplasmin | CP |
| 17455-42 | FOLR1 | FOLR1 |
| 17460-51 | Mx1 | MX1 |
| 17495-141 | SIR3 | SIRT3 |
| 17509-6 | HPPD | HPD |
| 17513-11 | ANX11 | ANXA11 |
| 17514-48 | RAB21 | RAB21 |
| 17672-184 | Gastric intrinsic factor | CBLIF |
| 17682-1 | CD46 | CD46 |
| 17685-9 | Apo A-IV | APOA4 |
| 17691-1 | TPP1 | TPP1 |
| 17702-53 | UGT 1A1 | UGT1A1 |
| 17706-4 | PPR1A | PPP1R1A |

| | | |
|---|---|---|
| 17721-82 | GAS7 | GAS7 |
| 17734-13 | a-endosulfine | ENSA |
| 17737-7 | IVD | IVD |
| 17739-1 | HCDH | HADH |
| 17742-2 | RRAS | RRAS |
| 17746-77 | FIS1 | FIS1 |
| 17747-45 | TRAF1 | TRAF1 |
| 17758-79 | DCXR | DCXR |
| 17764-108 | RHOC | RHOC |
| 17766-5 | NCF-1 | NCF1 |
| 17768-50 | HAOX1 | HAO1 |
| 17772-7 | UBD | UBD |
| 17783-9 | MMAB | MMAB |
| 17787-1 | Enoyl-CoA hydratase | ECHS1 |
| 17792-158 | SSDH | ALDH5A1 |
| 17794-6 | Phosphomannomutas | PMM2 |
| 17799-9 | 6PGL | PGLS |
| 17804-102 | PNPO | PNPO |
| 17843-30 | PPCS | PPCS |
| 17850-42 | KLF4 | KLF4 |
| 17857-6 | ACADL | ACADL |
| 18158-45 | Caspase-8 | CASP8 |
| 18162-167 | IRAK4 | IRAK4 |
| 18170-46 | P5CR1 | PYCR1 |
| 18182-24 | PCKGC | PCK1 |
| 18183-3 | ARH | LDLRAP1 |
| 18185-118 | ALDOB | ALDOB |
| 18187-16 | ACY2 | ASPA |
| 18188-12 | GATM | GATM |
| 18214-2 | GSH0 | GCLM |
| 18216-22 | IL-11 RA | IL11RA |
| 18226-148 | COX5A | COX5A |
| 18241-18 | HEM6 | CPOX |
| 18291-8 | p21 | CDKN1A |
| 18295-102 | GRHPR | GRHPR |
| 18297-8 | RASM | MRAS |
| 18309-18 | CISY | CS |
| 18319-7 | ODPX | PDHX |
| 18324-61 | SUOX | SUOX |
| 18331-3 | GLCM | GBA |
| 18347-15 | Laminin-2 | LAMC1\|LAMB1\|LAMA2 |
| 18376-19 | Myosin light chain 1 | MYL3 |
| 18380-78 | Albumin | ALB |
| 18381-16 | ALDH-E2 | ALDH2 |
| 18382-109 | Catechol O-methyltrar | COMT |
| 18396-10 | AES | TLE5 |
| 18398-1 | AK1D1 | AKR1D1 |

| | | |
|---|---|---|
| 18415-16 | ARL6 | ARL6 |
| 18429-10 | Cadherin E | CDH1 |
| 18830-1 | Omentin | ITLN1 |
| 18831-6 | LRIG1 | LRIG1 |
| 18870-1 | CD3E | CD3E |
| 18874-66 | CEBPA | CEBPA |
| 18880-81 | Collagen Type III | COL3A1 |
| 18890-227 | Fibrinogen B | FGB |
| 18891-98 | GBP2 | GBP2 |
| 18892-48 | GPC4 | GPC4 |
| 18897-31 | HDAC2 | HDAC2 |
| 18900-37 | H-ras (WT) | HRAS |
| 18901-26 | HS71B | HSPA1B |
| 18916-25 | Inosine triphosphatas | ITPA |
| 18921-30 | PHEX | PHEX |
| 18925-24 | Proteasome subunit al | PSMA5 |
| 18931-40 | SLIT3 | SLIT3 |
| 18934-50 | Tissue transglutamina | TGM2 |
| 18938-3 | VLDLR | VLDLR |
| 18950-13 | RAC2 | RAC2 |
| 19114-8 | KITM | TK2 |
| 19119-10 | DDIT3 | DDIT3 |
| 19120-33 | OXDA | DAO |
| 19121-3 | UBQL2 | UBQLN2 |
| 19127-1 | HSPB6 | HSPB6 |
| 19136-22 | MMSA | ALDH6A1 |
| 19144-9 | ITF2 | TCF4 |
| 19168-71 | PPIP1 | PSTPIP1 |
| 19176-27 | FA49B | CYRIB |
| 19187-21 | STABP | STAMBP |
| 19197-95 | THIL | ACAT1 |
| 19209-6 | IKB-alpha | NFKBIA |
| 19215-7 | NDRG1 | NDRG1 |
| 19230-12 | GSTT1 | GSTT1 |
| 19233-75 | ATOX1 | ATOX1 |
| 19238-12 | GLNA | GLUL |
| 19258-24 | GCDH | GCDH |
| 19262-219 | ACADV | ACADVL |
| 19273-3 | Glutathione reductase | GSR |
| 19274-80 | Alpha-1-syntrophin | SNTA1 |
| 19278-19 | RAB1B | RAB1B |
| 19289-29 | DCUP | UROD |
| 19290-5 | HPRT | HPRT1 |
| 19291-2 | CASQ2 | CASQ2 |
| 19293-6 | VP26A | VPS26A |
| 19296-51 | MLRA | MYL7 |
| 19297-4 | G6PD | G6PD |

| | | |
|---|---|---|
| 19303-64 | PT117 | PET117 |
| 19364-163 | PCNA | PCNA |
| 19365-11 | BCAT2 | BCAT2 |
| 19366-8 | PEX26 | PEX26 |
| 19561-216 | PLXD1 | PLXND1 |
| 19564-61 | IRF4 | IRF4 |
| 19568-17 | IL-15 | IL15 |
| 19570-12 | FGF-8 | FGF8 |
| 19602-36 | jun-D | JUND |
| 19637-9 | CRH | CRH |
| 19638-9 | Dynorphin A (1-17) | PDYN |
| 19639-53 | IAPP | IAPP |
| 19640-2 | Parathyroid Hormone: | PTH |
| 19751-21 | ADA | ADA |
| 19768-13 | Cystatin B | CSTB |
| 19823-75 | SIR1 | SIRT1 |
| 20073-22 | K-ras | KRAS |
| 20103-176 | N-glycosylase/DNA lya | OGG1 |
| 20107-11 | MYL4 | MYL4 |
| 20116-30 | OXSR1 | OXSR1 |
| 20128-1 | CREB1 | CREB1 |
| 20134-27 | MARCO | MARCO |
| 20165-4 | DPYS | DPYS |
| 20185-44 | ODP2 | DLAT |
| 20187-10 | Integrin aVb3 | ITGB3\|ITGAV |
| 20191-13 | Integrin aVb8 | ITGAV\|ITGB8 |
| 20195-13 | IFIH1 | IFIH1 |
| 20203-45 | C1s | C1S |
| 20211-75 | GBP5 | GBP5 |
| 20215-45 | Integrin aVb1 | ITGB1\|ITGAV |
| 20217-26 | Lamin-B2 | LMNB2 |
| 20225-119 | STA5A | STAT5A |
| 20241-9 | SNP23 | SNAP23 |
| 20243-26 | Profilin-1 | PFN1 |
| 20245-13 | Retinoic acid receptor | RXRA |
| 20374-41 | COQ9 | COQ9 |
| 20378-110 | LZTL1 | LZTFL1 |
| 20425-12 | HHEX | HHEX |
| 20428-5 | PHOP1 | PHOSPHO1 |
| 20458-22 | NDUA2 | NDUFA2 |
| 20516-11 | SCN3B | SCN3B |
| 20528-23 | CD68 | CD68 |
| 20533-39 | IL-35 | IL12A\|EBI3 |
| 20534-6 | SCRB1 | SCARB1 |
| 20542-47 | CALCR | CALCR |
| 20553-2 | P3IP1 | PIK3IP1 |
| 20564-53 | THY1 | THY1 |

| | | |
|---|---|---|
| 20572-6 | MER | MERTK |
| 20581-42 | GLP1R:ECD | GLP1R |
| 20590-13 | NPY | NPY |
| 20931-156 | AUHM | AUH |
| 20953-34 | MMAC | MMACHC |
| 20996-107 | SDHF1 | SDHAF1 |
| 21107-5 | CIA30 | NDUFAF1 |
| 21120-3 | MIMIT | NDUFAF2 |
| 21126-27 | GFPT1 | GFPT1 |
| 21207-1 | COASY | COASY |
| 21227-18 | SULT 1A1*2 | SULT1A1 |
| 21239-31 | CYH1 | CYTH1 |
| 21240-6 | GCST | AMT |
| 21247-16 | METK1 | MAT1A |
| 21269-198 | HYES | EPHX2 |
| 21271-53 | AK1C2 | AKR1C2 |
| 21280-13 | T22D3 | TSC22D3 |
| 21330-13 | SYYM | YARS2 |
| 21355-4 | USF1 | USF1 |
| 21384-2 | Alpha-L-fucosidase I | FUCA1 |
| 21393-62 | SAHH | AHCY |
| 21430-4 | cPLA2-alpha | PLA2G4A |
| 21441-20 | gp75 | TYRP1 |
| 21452-3 | DNM1L | DNM1L |
| 21477-105 | MAEA | MAEA |
| 21478-20 | SMAD5 | SMAD5 |
| 21483-155 | KAP0 | PRKAR1A |
| 21487-20 | BL1S6 | BLOC1S6 |
| 21491-7 | VAP-1 | AOC3 |
| 21510-24 | GRK5 | GRK5 |
| 21513-1 | Caspase-7 | CASP7 |
| 21514-1 | GP1BB | GP1BB |
| 21533-51 | LSP1 | LSP1 |
| 21552-9 | ACOX1 | ACOX1 |
| 21572-91 | XYLT2 | XYLT2 |
| 21579-35 | MYPC3 | MYBPC3 |
| 21599-6 | CGL | CTH |
| 21643-8 | RS20 | RPS20 |
| 21651-9 | TXN4A | TXNL4A |
| 21660-4 | SAR1B | SAR1B |
| 21685-29 | DCC | DCC |
| 21703-31 | Activin RIA | ACVR1 |
| 21705-33 | METRL | METRNL |
| 21724-22 | GALNS | GALNS |
| 21739-7 | GLCNE | GNE |
| 21766-50 | SPHM | SGSH |
| 21796-43 | Glutathione peroxidas | GPX3 |

| | | |
|---|---|---|
| 21802-53 | NEIL1 | NEIL1 |
| 21814-13 | MMAD | MMADHC |
| 21817-5 | MYL9 | MYL9 |
| 21833-6 | sirtuin | SIRT6 |
| 21856-59 | KHK | KHK |
| 21875-31 | ZFAN3 | ZFAND3 |
| 21901-14 | Integrin a3b1 | F11 |
| 21903-6 | Integrin aLb2 | ITGA3\|ITGB1 |
| 21905-10 | Integrin alpha-2/ b1 | ITGB2\|ITGAL |
| 2190-55 | Coagulation Factor XI | ITGB1\|ITGA2 |
| 21909-2 | Integrin a5b1 | ITGA5\|ITGB1 |
| 21911-17 | NDP | NDP |
| 21967-20 | DHI1 | HSD11B1 |
| 21969-5 | DPP6 | DPP6 |
| 21976-4 | IKK-gamma | IKBKG |
| 21981-2 | Integrin alpha-M | ITGAM |
| 21987-76 | LPL | LPL |
| 21995-20 | NOS | NOS3 |
| 22003-4 | Paraoxonase-2 | PON2 |
| 22005-8 | PDP1 | PDP1 |
| 22009-1 | PPM1B | PPM1B |
| 2201-17 | Endostatin | COL18A1 |
| 22019-21 | SH2B3 | SH2B3 |
| 22043-174 | HSP70 protein 2 | HSPA2 |
| 22045-8 | SUMO4 | SUMO4 |
| 22047-46 | CO5A1 | COL5A1 |
| 22049-24 | 14-3-3 eta | YWHAH |
| 22057-9 | AL1B1 | ALDH1B1 |
| 22075-16 | ATF3 | ATF3 |
| 22078-4 | ATTY | TAT |
| 22088-3 | BTG1 | BTG1 |
| 22091-14 | BUP1 | UPB1 |
| 22116-9 | CREM | CREM |
| 2212-69 | tPA | PLAT |
| 22148-135 | FOXP3 | FOXP3 |
| 22154-37 | GATD1 | GATAD1 |
| 22383-21 | MITF | MITF |
| 22394-8 | TCF21 | TCF21 |
| 22395-7 | Tristetraproline | ZFP36 |
| 22397-2 | XBP1 | XBP1 |
| 22405-61 | PCKGM | PCK2 |
| 22485-1 | KCTD7 | KCTD7 |
| 22488-17 | KLF9 | KLF9 |
| 22505-24 | NAKD2 | NADK2 |
| 22508-2 | NDUBA | NDUFB10 |
| 22509-9 | NDUF5 | NDUFAF5 |
| 22510-6 | NF2L2 | NFE2L2 |

| | | |
|---|---|---|
| 22526-81 | PDHB | PDHB |
| 22527-4 | PDLI3 | PDLIM3 |
| 22529-31 | PHF6 | PHF6 |
| 22531-44 | PITX2 | PITX2 |
| 22540-129 | PSB7 | PSMB7 |
| 22544-10 | RA51C | RAD51C |
| 22569-55 | TGFR-1 | TGFBR1 |
| 22584-2 | LRP6 | LRP6 |
| 22773-69 | SDHB | SDHB |
| 22776-40 | SHP | NR0B2 |
| 2278-61 | TIMP-2 | TIMP2 |
| 22800-24 | TELT | TCAP |
| 22808-59 | THB | THRB |
| 22818-4 | TXTP | SLC25A1 |
| 22821-50 | UNG | UNG |
| 22847-18 | STK4 | STK4 |
| 22953-85 | CEBPB | CEBPB |
| 22958-6 | DEST | DSTN |
| 22969-12 | MCP-3 | CCL7 |
| 22978-13 | HMG-2 | HMGB2 |
| 23022-5 | RhoGDI | GDI1 |
| 23024-25 | SAR1A | SAR1A |
| 23038-63 | UNC5B | UNC5B |
| 23039-56 | MEOX2 | MEOX2 |
| 23173-3 | TIMP-1 | TIMP1 |
| 23213-21 | CK070 | CFAP300 |
| 23254-31 | NDUF3 | NDUFAF3 |
| 23263-9 | KCD15 | KCTD15 |
| 23275-9 | SSPN | SSPN |
| 23294-19 | MLX | MLX |
| 2330-2 | SDF-1 | CXCL12 |
| 23302-19 | AKTS1 | AKT1S1 |
| 23314-46 | CC103 | CCDC103 |
| 2333-72 | TGF-b1 | TGFB1 |
| 23352-9 | DTBP1 | DTNBP1 |
| 23369-17 | PAHX | PHYH |
| 23393-56 | ABHD5 | ABHD5 |
| 23404-16 | MD2L2 | MAD2L2 |
| 23418-66 | CCD92 | CCDC92 |
| 23555-11 | RAD | RRAD |
| 23557-110 | CAPZB | CAPZB |
| 23567-37 | GNAQ | GNAQ |
| 23568-41 | MEIS2 | MEIS2 |
| 23643-22 | DHB4 | HSD17B4 |
| 23652-15 | SOX | PIPOX |
| 23654-6 | ALG2 | ALG2 |
| 23658-1 | SYNM | NARS2 |

| | | |
|---|---|---|
| 23666-35 | PUR9 | ATIC |
| 23672-10 | PSMD6 | PSMD6 |
| 23673-9 | ODB2 | DBT |
| 23680-1 | MCPIP | ZC3H12A |
| 23694-3 | ASNS | ASNS |
| 23703-8 | PDLI5 | PDLIM5 |
| 23903-3 | GOT2 | GOT2 |
| 23915-18 | Rab-7 | RAB7A |
| 23981-172 | CBPA1 | CPA1 |
| 24023-35 | sEPCR | PROCR |
| 24050-26 | GSK-3 beta | GSK3B |
| 2418-55 | Apo E | APOE |
| 24211-25 | HMGCL | HMGCL |
| 24215-8 | PRRX1 | PRRX1 |
| 24221-3 | PPAR gamma | PPARG |
| 24223-5 | GST2 | GSTT2 |
| 24252-85 | CL065 | MTRFR |
| 24263-6 | OCLN | OCLN |
| 24276-171 | TTC25 | ODAD4 |
| 24278-4 | UBP3 | USP3 |
| 2436-49 | CXCL16, soluble | CXCL16 |
| 24407-31 | KIF3C | KIF3C |
| 24414-3 | Glycogen phosphoryla | PYGB |
| 24419-3 | MOCOS | MOCOS |
| 24426-15 | CTNA1 | CTNNA1 |
| 24459-15 | COPB2 | COPB2 |
| 24463-1 | NDRG2 | NDRG2 |
| 24469-10 | LPIN1 | LPIN1 |
| 24487-95 | BL1S3 | BLOC1S3 |
| 24646-47 | SPYA | AGXT |
| 24651-1 | RTN1 | RTN1 |
| 24659-6 | DJB13 | DNAJB13 |
| 24671-15 | ASC | PYCARD |
| 24676-105 | CU059 | CFAP298 |
| 24681-2 | SERB | CCL20 |
| 24682-35 | CYBR1 | PSPH |
| 2468-62 | MIP-3a | CYBRD1 |
| 24689-9 | PTRF | CAVIN1 |
| 24694-158 | NHERF | SLC9A3R1 |
| 24701-21 | DTNA | DTNA |
| 2474-54 | SAP | APCS |
| 2475-1 | SCF sR | KIT |
| 24898-39 | SCFD1 | SCFD1 |
| 24902-84 | GEPH | GPHN |
| 24910-18 | NonO protein | NONO |
| 24940-3 | IKK-beta | IKBKB |
| 24958-3 | RFX5 | RFX5 |

| | | |
|---|---|---|
| 24960-48 | HIP1R | HIP1R |
| 25033-194 | TER ATPase | VCP |
| 25043-74 | SCOT | OXCT1 |
| 25051-104 | FTO | FTO |
| 25060-18 | MAOX | ME1 |
| 25061-8 | ELMO2 | ELMO2 |
| 25066-32 | GLYG2 | GYG2 |
| 25076-2 | Selenium-binding prot | SELENBP1 |
| 25088-42 | PHKA1 | PHKA1 |
| 25089-21 | KRIP-1 | TRIM28 |
| 25100-11 | CMIP | CMIP |
| 25105-70 | EIF2A | EIF2A |
| 25209-15 | ATNG | FXYD2 |
| 25233-2 | MPI | MPI |
| 2524-56 | HMG-1 | HMGB1 |
| 25253-17 | GNAS | GNAS |
| 25264-102 | SHIP | INPP5D |
| 25283-2 | CTNA3 | CTNNA3 |
| 25288-16 | FMO3 | FMO3 |
| 25291-27 | PKHM2 | PLEKHM2 |
| 25292-4 | EST3 | CES3 |
| 25297-11 | SPD2B | SH3PXD2B |
| 25300-39 | MALT1 | MALT1 |
| 25308-8 | LIMK1 | LIMK1 |
| 25460-36 | IRF5 | IRF5 |
| 25463-3 | KAP3 | PRKAR2B |
| 25464-1 | CP2CJ | CYP2C19 |
| 2570-72 | IGFBP-2 | IGFBP2 |
| 2571-12 | IGFBP-3 | IGFBP3 |
| 2578-67 | MCP-1 | CCL2 |
| 2579-17 | MMP-9 | MMP9 |
| 2580-83 | Myeloperoxidase | MPO |
| 2585-2 | PRL | PRL |
| 25880-14 | 5-Lipoxygenase | ALOX5 |
| 25886-11 | ABCD4 | ABCD4 |
| 25902-27 | DHE3 | GLUD1 |
| 25913-17 | FRIH | FTH1 |
| 25917-12 | Hexosaminidase A | HEXA |
| 25922-7 | LRP5 | LRP5 |
| 25938-13 | NGN3 | NEUROG3 |
| 25951-17 | PRAME | PRAME |
| 25962-51 | WNK1 | WNK1 |
| 2597-8 | VEGF | VEGFA |
| 2609-59 | Cystatin C | CST3 |
| 2611-72 | Dtk | TYRO3 |
| 2615-60 | Ephrin-A5 | EFNA5 |
| 2618-10 | ERBB4 | ERBB4 |

| | | |
|---|---|---|
| 2619-72 | GA733-1 protein | TACSTD2 |
| 2620-4 | gp130, soluble | IL6ST |
| 2622-18 | HO-2 | HMOX2 |
| 2631-50 | IL-10 Rb | IL10RB |
| 2632-5 | IL-12 Rb1 | IL12RB1 |
| 2633-52 | IL-13 Ra1 | IL13RA1 |
| 2634-2 | IL-2 sRg | IL2RG |
| 2636-10 | Lymphotoxin b R | LTBR |
| 2637-77 | Macrophage mannose | MRC1 |
| 2644-11 | PKC-A | PRKCA |
| 2645-54 | PKC-Z | PRKCZ |
| 2649-77 | sICAM-3 | ICAM3 |
| 2652-15 | suPAR | PLAUR |
| 2654-19 | TNF sR-I | TNFRSF1A |
| 2666-53 | Bone proteoglycan II | DCN |
| 2670-67 | CK-MM | CKM |
| 2677-1 | ERBB1 | EGFR |
| 2681-23 | HGF | HGF |
| 2682-68 | HSP 60 | HSPD1 |
| 2686-67 | IGFBP-6 | IGFBP6 |
| 2692-74 | NPS-PLA2 | PLA2G2A |
| 2695-25 | PECAM-1 | PECAM1 |
| 2697-7 | PF-4 | PF4 |
| 2700-56 | Protein S | PROS1 |
| 2704-74 | TACI | TNFRSF13B |
| 2706-69 | Thyroxine-Binding Glo | SERPINA7 |
| 2730-58 | MICA | MICA |
| 2731-29 | NADPH-P450 Oxidored | POR |
| 2750-3 | Apo A-I | APOA1 |
| 2753-2 | C1q | C1QC\|C1QB\|C1QA |
| 2754-50 | C3 | C3 |
| 2765-4 | GDF-11/8 | MSTN\|GDF11 |
| 2768-56 | Hemopexin | HPX |
| 2773-50 | IL-10 | IL10 |
| 2774-10 | IL-16 | IL16 |
| 2778-10 | IL-22 | IL22 |
| 2780-35 | Lactoferrin | LTF |
| 2788-55 | MMP-3 | MMP3 |
| 2789-26 | MMP-7 | MMP7 |
| 2790-54 | NAP-2 | PPBP |
| 2794-60 | SOD | SOD1 |
| 2796-62 | Fibrinogen | FGG\|FGB\|FGA |
| 2797-56 | Apo B | APOB |
| 2805-6 | ACE2 | ACE2 |
| 2811-27 | Angiopoietin-1 | ANGPT1 |
| 2813-11 | ART | AGRP |
| 2819-23 | Cadherin-5 | CDH5 |

| | | |
|---|---|---|
| 2827-23 | Fractalkine/CX3CL-1 | CX3CL1 |
| 2829-19 | IL-27 | EBI3\|IL27 |
| 2831-29 | Kallikrein 11 | KLK11 |
| 2834-54 | kallikrein 8 | KLK8 |
| 2836-68 | Lipocalin 2 | LCN2 |
| 2843-13 | SPINT2 | SPINT2 |
| 2855-49 | ERK-1 | MAPK3 |
| 2857-70 | Glucocorticoid recepto | NR3C1 |
| 2859-69 | HDAC8 | HDAC8 |
| 2864-2 | MEK1 | MAP2K1 |
| 2867-52 | PKB | AKT1 |
| 2869-68 | PKC-D | PRKCD |
| 2870-29 | RAC1 | RAC1 |
| 2871-73 | RAD51 | RAD51 |
| 2878-66 | YES | YES1 |
| 2900-53 | HCC-1 | CCL14 |
| 2906-55 | IL-4 | IL4 |
| 2925-9 | PAI-1 | SERPINE1 |
| 2942-50 | Cytochrome c | CYCS |
| 2943-5 | Cytochrome P450 3A4 | CYP3A4 |
| 2948-58 | Growth hormone rece | GHR |
| 2953-31 | Luteinizing hormone | LHB\|CGA |
| 2961-1 | Protein C | PROC |
| 2962-50 | PTHrP | PTHLH |
| 2967-8 | VCAM-1 | VCAM1 |
| 2970-60 | AREG | AREG |
| 2973-15 | CD36 ANTIGEN | CD36 |
| 2974-61 | contactin-1 | CNTN1 |
| 2975-19 | CTGF | CCN2 |
| 2976-58 | Desmoglein-1 | DSG1 |
| 2985-35 | Gro-a | CXCL1 |
| 2986-49 | Gro-g | CXCL3 |
| 2991-9 | IL-1 sRI | IL1R1 |
| 2992-59 | IL-17 sR | IL17RA |
| 2997-8 | JAM-B | JAM2 |
| 3000-66 | MBL | MBL2 |
| 3005-5 | PTP-1B | PTPN1 |
| 3009-3 | TGF-b R III | TGFBR3 |
| 3022-4 | CTLA-4 | CTLA4 |
| 3025-50 | bFGF | FGF2 |
| 3032-11 | FSH | FSHB\|CGA |
| 3033-57 | Galectin-2 | LGALS2 |
| 3034-1 | GFAP | GFAP |
| 3035-80 | IL-19 | IL19 |
| 3037-62 | IL-1b | IL1B |
| 3038-9 | I-TAC | CXCL11 |
| 3040-59 | MIP-1a | CCL3 |

| | | |
|---|---|---|
| 3044-3 | PARC | CCL18 |
| 3046-31 | resistin | RETN |
| 3049-61 | Trypsin | PRSS1 |
| 3050-7 | vWF | VWF |
| 3052-8 | Fas ligand, soluble | FASLG |
| 3054-3 | Haptoglobin, Mixed Ty | HP |
| 3055-54 | IL-4 sR | IL4R |
| 3059-50 | BAFF | TNFSF13B |
| 3065-65 | FGF-5 | FGF5 |
| 3066-12 | Galectin-3 | LGALS3 |
| 3070-1 | IL-2 | IL2 |
| 3073-51 | IL-18 BPa | IL18BP |
| 3074-6 | LBP | LBP |
| 3077-66 | Coagulation Factor Xa | F10 |
| 3078-1 | PlGF | PGF |
| 3079-62 | TIG2 | RARRES2 |
| 3115-64 | MK01 | MAPK1 |
| 3122-6 | SMAC | DIABLO |
| 3143-3 | sCD4 | CD4 |
| 3148-49 | Gro-b | CXCL2 |
| 3151-6 | IL-2 sRa | IL2RA |
| 3169-70 | IDUA | IDUA |
| 3171-57 | amyloid precursor pro | APP |
| 3172-28 | ARSB | ARSB |
| 3175-51 | ATS13 | ADAMTS13 |
| 3179-51 | Cathepsin A | CTSA |
| 3181-50 | Cathepsin S | CTSS |
| 3184-25 | Coagulation Factor VII | F7 |
| 3186-2 | C2 | C2 |
| 3189-61 | Enterokinase | TMPRSS15 |
| 3198-4 | IDS | IDS |
| 3204-2 | LKHA4 | LTA4H |
| 3206-4 | LYVE1 | LYVE1 |
| 3209-69 | MEPE | MEPE |
| 3212-30 | ASAH2 | ASAH2 |
| 3213-65 | Nidogen | NID1 |
| 3220-40 | RET | RET |
| 3221-54 | SARP-2 | SFRP1 |
| 3232-28 | TrATPase | ACP5 |
| 3283-21 | BGH3 | TGFBI |
| 3285-23 | C1r | C1R |
| 3294-55 | CFC1 | CFC1 |
| 3309-2 | FCG2A | FCGR2A |
| 3310-62 | FCG2B | FCGR2B |
| 3311-27 | FCG3B | FCGR3B |
| 3312-64 | FCGR1 | FCGR1A |
| 3316-58 | Heparin cofactor II | SERPIND1 |

| | | |
|---|---|---|
| 3317-33 | HTRA2 | HTRA2 |
| 3320-49 | IGFBP-7 | IGFBP7 |
| 3323-37 | LRP8 | LRP8 |
| 3327-27 | NET4 | NTN4 |
| 3332-57 | RGM-C | HJV |
| 3336-50 | TFPI | TFPI |
| 3339-33 | TSP2 | THBS2 |
| 3341-33 | ABL1 | ABL1 |
| 3343-1 | Aminoacylase-1 | ACY1 |
| 3344-60 | Antithrombin III | SERPINC1 |
| 3347-9 | BARK1 | GRK2 |
| 3350-53 | CAMK2A | CAMK2A |
| 3359-11 | CDK8/cyclin C | CDK8\|CCNC |
| 3360-50 | Chk2 | CHEK2 |
| 3367-8 | FETUB | FETUB |
| 3389-7 | PCI | SERPINA5 |
| 3390-72 | PIK3CA/PIK3R1 | PIK3R1\|PIK3CA |
| 3391-10 | PK3CG | PIK3CG |
| 3396-54 | Renin | REN |
| 3400-49 | TBK1 | TBK1 |
| 3412-7 | Bcl-2 | BCL2 |
| 3419-49 | CAMK2D | CAMK2D |
| 3423-59 | Chymase | CMA1 |
| 3432-21 | EPHA3 | EPHA3 |
| 3435-53 | FN1.4 | FN1 |
| 3437-80 | Flt-3 | FLT3 |
| 3447-64 | IL-8 | CXCL8 |
| 3448-13 | IR | INSR |
| 3452-17 | LCK | LCK |
| 3459-49 | PDGF Rb | PDGFRB |
| 3470-1 | sE-Selectin | SELE |
| 3474-19 | Thrombospondin-1 | THBS1 |
| 3484-60 | Angiotensinogen | AGT |
| 3486-58 | b-ECGF | FGF1 |
| 3488-64 | Catalase | CAT |
| 3489-9 | CNTF | CNTF |
| 3497-13 | IFN-aA | IFNA2 |
| 3498-53 | IL-17 | IL17A |
| 3503-4 | Integrin a1b1 | ITGB1\|ITGA1 |
| 3504-58 | LEAP-1 | HAMP |
| 3505-6 | Lymphotoxin a1/b2 | LTA\|LTB |
| 3514-49 | Proteinase-3 | PRTN3 |
| 3518-54 | TAFI | CPB2 |
| 3519-3 | TARC | CCL17 |
| 3520-58 | TGF-b3 | TGFB3 |
| 3521-16 | TSH | TSHB\|CGA |
| 3522-57 | Vasoactive Intestinal P | VIP |

| | | |
|---|---|---|
| 3534-14 | CD40 ligand, soluble | CD40LG |
| 3535-84 | DKK1 | DKK1 |
| 3538-26 | dopa decarboxylase | DDC |
| 3554-24 | Adiponectin | ADIPOQ |
| 3580-25 | a1-Antitrypsin | SERPINA1 |
| 3585-54 | BASI | BSG |
| 3593-72 | Caspase-3 | CASP3 |
| 3600-2 | Chitotriosidase-1 | CHIT1 |
| 3601-54 | CHL1 | CHL1 |
| 3605-77 | MASP3:Light | MASP1 |
| 3611-70 | Endothelin-converting | ECE1 |
| 3623-84 | LY86 | LY86 |
| 3628-3 | MP2K2 | MAP2K2 |
| 3642-4 | SLAF5 | CD84 |
| 3647-49 | TLR4:MD-2 complex | TLR4\|LY96 |
| 3651-50 | VEGF sR2 | KDR |
| 3666-17 | complement factor H-r | CFHR5 |
| 3708-62 | a2-Macroglobulin | A2M |
| 3709-4 | ALT | GPT |
| 3710-49 | Angiostatin | PLG |
| 3714-49 | CK-MB | CKM\|CKB |
| 3719-2 | p27Kip1 | CDKN1B |
| 3727-35 | PYY | PYY |
| 3728-52 | Secretin | SCT |
| 3730-81 | TNR4 | TNFRSF4 |
| 3736-60 | BMP-6 | BMP6 |
| 3739-72 | gpIIbIIIa | ITGB3\|ITGA2B |
| 3758-63 | Activated Protein C | PROC |
| 3761-4 | COX-2 | PTGS2 |
| 3773-15 | sTie-2 | TEK |
| 3796-79 | ANGL4 | ANGPTL4 |
| 3797-1 | Cadherin-2 | CDH2 |
| 3799-11 | Carbonic anhydrase III | CA3 |
| 3800-71 | CK-BB | CKB |
| 3807-1 | FGF23 | FGF23 |
| 3808-76 | FGFR-2 | FGFR2 |
| 3809-1 | FGFR-3:CD | FGFR3 |
| 3817-18 | KPCT | PRKCQ |
| 3825-18 | MK08 | MAPK8 |
| 3831-21 | pTEN | PTEN |
| 3835-11 | TLR2 | TLR2 |
| 3837-6 | ZAP70 | ZAP70 |
| 3847-56 | ETHE1 | ETHE1 |
| 3848-14 | GAPDH, liver | GAPDH |
| 3853-56 | MDHC | MDH1 |
| 3854-24 | NACA | NACA |
| 3855-56 | Peroxiredoxin-1 | PRDX1 |

| | | |
|---|---|---|
| 3860-7 | PSA6 | PSMA6 |
| 3881-49 | DLC8 | DYNLL1 |
| 3889-64 | Lamin-B1 | LMNB1 |
| 3890-8 | LDH-H 1 | LDHB |
| 4125-52 | sRAGE | AGER |
| 4127-75 | C6 | C6 |
| 4128-27 | Eotaxin-2 | CCL24 |
| 4130-71 | FGF-6 | FGF6 |
| 4131-72 | Fibronectin | FN1 |
| 4132-27 | FST | FST |
| 4138-25 | IL-20 | IL20 |
| 4139-71 | IL-6 sRa | IL6R |
| 4141-79 | IP-10 | CXCL10 |
| 4148-49 | PAPP-A | PAPPA |
| 4149-8 | PDGF-BB | PDGFB |
| 4150-75 | Plasmin | PLG |
| 4151-6 | Plasminogen | PLG |
| 4152-58 | Prekallikrein | KLKB1 |
| 4153-11 | alpha-1-antichymotryp | KLK3\|SERPINA3 |
| 4154-57 | P-Selectin | SELP |
| 4156-74 | TGF-b2 | TGFB2 |
| 4157-2 | Thrombin | F2 |
| 4158-54 | uPA | PLAU |
| 4160-49 | MMP-2 | MMP2 |
| 4162-54 | Transferrin | TF |
| 4232-19 | IGF-I sR | IGF1R |
| 4234-8 | IL-1 R4 | IL1RL1 |
| 4240-31 | M2-PK | PKM |
| 4245-80 | MDM2 | MDM2 |
| 4261-55 | paraoxonase 1 | PON1 |
| 4276-10 | prostatic binding prote | PEBP1 |
| 4280-47 | PSA2 | PSMA2 |
| 4306-4 | Transketolase | TKT |
| 4309-59 | Triosephosphate isom | TPI1 |
| 4318-12 | PTP-1C | PTPN6 |
| 4322-28 | AMNLS | AMN |
| 4328-2 | BOC | BOC |
| 4337-49 | CRP | CRP |
| 4342-10 | sICAM-1 | ICAM1 |
| 4374-45 | MIC-1 | GDF15 |
| 4413-3 | SLPI | SLPI |
| 4414-69 | SP-D | SFTPD |
| 4423-77 | BCL2-like 1 protein | BCL2L1 |
| 4430-44 | Collectin Kidney 1 | COLEC11 |
| 4455-89 | MFGM | MFGE8 |
| 4460-8 | PDPK1 | PDPK1 |
| 4467-49 | SPARCL1 | SPARCL1 |

| | | |
|---|---|---|
| 4468-21 | SPHK2 | SPHK2 |
| 4479-14 | C1-Esterase Inhibitor | SERPING1 |
| 4481-34 | C4 | C4B\|C4A |
| 4482-66 | C5b, 6 Complex | C6\|C5 |
| 4493-92 | IL-11 | IL11 |
| 4496-60 | MMP-12 | MMP12 |
| 4498-62 | NCAM-120 | NCAM1 |
| 4499-21 | PDGF-AA | PDGFA |
| 4500-50 | SCGF-alpha | CLEC11A |
| 4541-49 | CDON | CDON |
| 4542-24 | Clusterin | CLU |
| 4545-53 | DnaJ homolog | DNAJC19 |
| 4546-27 | EMR2 | ADGRE2 |
| 4559-64 | KYNU | KYNU |
| 4673-13 | IL-6 | IL6 |
| 4697-59 | GM-CSF | CSF2 |
| 4703-87 | TNF-b | LTA |
| 4719-58 | Protein disulfide isome | PDIA3 |
| 4721-54 | TFF3 | TFF3 |
| 4775-34 | Gelsolin | GSN |
| 4807-13 | CO8A1 | COL8A1 |
| 4831-4 | sL-Selectin | SELL |
| 4832-75 | TRAIL R1 | TNFRSF10A |
| 4834-61 | Epithelial cell kinase | EPHA2 |
| 4842-62 | Glypican 3 | GPC3 |
| 4851-25 | IL-1a | IL1A |
| 4859-6 | BMPR1A | BMPR1A |
| 4862-63 | BMP RII | BMPR2 |
| 4866-59 | TrkB | NTRK2 |
| 4867-15 | VEGF121 | VEGFA |
| 4874-3 | Angiogenin | ANG |
| 4876-32 | Coagulation Factor IX | F9 |
| 4880-21 | GDF2 | GDF2 |
| 4883-56 | Insulin | INS |
| 4889-82 | WNT7A | WNT7A |
| 4890-10 | ACTH | POMC |
| 4891-50 | Glucagon | GCG |
| 4903-72 | Calcineurin | PPP3R1\|PPP3CA |
| 4906-35 | Coagulation Factor V | F5 |
| 4908-6 | Endoglin | ENG |
| 4914-10 | HCG | CGB7\|CGB3\|CGA |
| 4915-64 | Hemoglobin | HBB\|HBA1 |
| 4917-62 | Integrin aVb5 | ITGB5\|ITGAV |
| 4920-10 | Lysozyme | LYZ |
| 4924-32 | MMP-1 | MMP1 |
| 4929-55 | SHBG | SHBG |
| 4931-59 | TF | F3 |

| | | |
|---|---|---|
| 4956-2 | EPI | EREG |
| 4960-72 | annexin I | ANXA1 |
| 4964-67 | ARTS1 | ERAP1 |
| 4969-2 | Carbonic anhydrase I | CA1 |
| 4970-55 | carbonic anhydrase II | CA2 |
| 4971-1 | CATZ | CTSZ |
| 4973-18 | cIAP-2 | BIRC3 |
| 4976-57 | CRK | CRK |
| 4979-34 | DERM | DPT |
| 4982-54 | Elafin | PI3 |
| 4985-11 | FABPE | FABP5 |
| 4986-59 | FAK1 | PTK2 |
| 4990-87 | GP1BA | GP1BA |
| 4992-49 | GRN | GRN |
| 5002-76 | MMP-14 | MMP14 |
| 5004-69 | MK11 | MAPK11 |
| 5007-1 | MAPK14 | MAPK14 |
| 5008-51 | Mn SOD | SOD2 |
| 5009-11 | Moesin | MSN |
| 5011-11 | PBEF | NAMPT |
| 5012-67 | Myokinase, human | AK1 |
| 5015-15 | PAFAH | PLA2G7 |
| 5018-68 | Peroxiredoxin-6 | PRDX6 |
| 5020-50 | phosphoglycerate kina | PGK1 |
| 5023-23 | PUR8 | ADSL |
| 5024-67 | Rb | RB1 |
| 5028-59 | sCD163 | CD163 |
| 5033-27 | Tropomyosin 1 alpha c | TPM1 |
| 5034-79 | Trypsin 2 | PRSS2 |
| 5036-50 | TSG-6 | TNFAIP6 |
| 5061-27 | B7-H2 | ICOSLG |
| 5080-131 | GPNMB:ECD | GPNMB |
| 5088-175 | IL-23 R | IL23R |
| 5089-11 | IL-7 Ra | IL7R |
| 5090-49 | ILT-2 | LILRB1 |
| 5092-51 | JAG1:ECD | JAG1 |
| 5100-53 | LIMP II | SCARB2 |
| 5107-7 | Notch 1 | NOTCH1 |
| 5108-72 | Notch-3 | NOTCH3 |
| 5111-15 | NRX3B | NRXN3 |
| 5117-14 | ROBO3 | ROBO3 |
| 5133-17 | TGF-b R II | TGFBR2 |
| 5138-50 | TWEAKR | TNFRSF12A |
| 5183-53 | AMPK a1b1g1 | PRKAG1\|PRKAB1\|PRKAA1 |
| 5223-59 | GCKR | GCKR |
| 5228-25 | KIF23 | KIF23 |
| 5230-99 | HMGR | HMGCR |

| | | |
|---|---|---|
| 5245-40 | AMPK a2b2g1 | PRKAG1\|PRKAB2\|PRKAA2 |
| 5254-69 | PDE3A | PDE3A |
| 5259-2 | TAK1-TAB1 | TAB1\|MAP3K7 |
| 5260-80 | TYK2 | TYK2 |
| 5264-65 | calreticulin | CALR |
| 5272-55 | SHC1:SH2 | SHC1 |
| 5301-7 | Eotaxin | CCL11 |
| 5312-49 | Apo E2 | APOE |
| 5315-22 | Troponin T | TNNT2 |
| 5316-54 | Prothrombin | F2 |
| 5350-14 | GPC6 | GPC6 |
| 5351-52 | hnRNP A2/B1 | HNRNPA2B1 |
| 5352-11 | HVEM | TNFRSF14 |
| 5353-89 | IL-1Ra | IL1RN |
| 5360-9 | PKB beta | AKT2 |
| 5400-52 | sLeptin R | LEPR |
| 5412-53 | CD27 | CD27 |
| 5424-55 | RANK | TNFRSF11A |
| 5430-66 | SHPS1 | SIRPA |
| 5441-67 | Troponin I | TNNI3 |
| 5443-62 | ANP | NPPA |
| 5451-1 | ALCAM | ALCAM |
| 5468-67 | IL-17 RC | IL17RC |
| 5475-10 | PKC-B-II | PRKCB |
| 5480-49 | RANTES | CCL5 |
| 5481-16 | RASA1 | RASA1 |
| 5508-62 | Cathepsin D | CTSD |
| 5509-7 | EGF:ECD | EGF |
| 5532-53 | bFGF-R | FGFR1 |
| 5534-49 | TRAIL R2:ECD | TNFRSF10B |
| 5542-22 | NRP1 | NRP1 |
| 5584-21 | Holo-TC II | TCN2 |
| 5620-13 | AMD | PAM |
| 5621-64 | THSD1 | THSD1 |
| 5626-20 | CTRC | CTRC |
| 5627-53 | GON1 | GNRH1 |
| 5635-66 | TREM2 | TREM2 |
| 5658-64 | coagulation factor XIII | F13B |
| 5659-11 | TRH | TRH |
| 5660-51 | SOD3 | SOD3 |
| 5661-15 | IL-18 | IL18 |
| 5666-64 | IL-1 sRII | IL1R2 |
| 5675-6 | Tenascin | TNC |
| 5682-13 | VASN | VASN |
| 5692-79 | TNF-a | TNF |
| 5698-60 | Tenascin-X | TNXB |
| 5707-55 | ELK3 | ELK3 |

| | | |
|---|---|---|
| 5743-82 | CART | CARTPT |
| 5746-37 | EFNB1 | EFNB1 |
| 5748-20 | Acid ceramidase | ASAH1 |
| 5757-45 | GNAS3 | GNAS |
| 5792-8 | AFP | AFP |
| 5801-72 | b-NGF | NGF |
| 5807-77 | CD70 | CD70 |
| 5810-25 | Cripto | TDGF1 |
| 5813-58 | Epo | EPO |
| 5825-49 | IFN-g R1 | IFNGR1 |
| 5854-60 | tau | MAPT |
| 5864-10 | aldolase A | ALDOA |
| 5867-60 | ARGI1 | ARG1 |
| 5870-23 | BAD | BAD |
| 5903-91 | HSP70 protein 8 | HSPA8 |
| 5909-51 | Nucleoside diphosphat | NME1 |
| 5915-58 | PEX5 | PEX5 |
| 5934-1 | Ferritin | FTL\|FTH1 |
| 5939-42 | TWEAK | TNFSF12 |
| 5954-62 | PTH | PTH |
| 5957-30 | Somatostatin-28 | SST |
| 5980-55 | BOLA3 | BOLA3 |
| 6003-28 | NEUR1 | NEU1 |
| 6020-52 | Urotensin-II | UTS2 |
| 6024-68 | CBPE | CPE |
| 6027-31 | REL3 | RLN3 |
| 6035-2 | SIAT1 | ST6GAL1 |
| 6049-64 | PTPRS | PTPRS |
| 6077-63 | CECR1 | ADA2 |
| 6123-69 | p53 | TP53 |
| 6207-10 | prosaposin | PSAP |
| 6232-54 | B7-2 | CD86 |
| 6245-4 | Nectin-2 | NECTIN2 |
| 6350-43 | Apo C-II | APOC2 |
| 6373-54 | DLK1 | DLK1 |
| 6375-75 | XXLT1 | XXYLT1 |
| 6379-62 | ATL2 | ADAMTSL2 |
| 6382-17 | MANBA | MANBA |
| 6383-90 | TLL1 | TLL1 |
| 6398-12 | LONM | LONP1 |
| 6420-4 | SDHF2 | SDHAF2 |
| 6433-57 | FA20A | FAM20A |
| 6440-31 | MFAP5 | MFAP5 |
| 6447-73 | PTX3 | PTX3 |
| 6448-36 | Sema E | SEMA3C |
| 6461-54 | Apo C-III | APOC3 |
| 6462-12 | TIMP-4 | TIMP4 |

| | | |
|---|---|---|
| 6495-14 | Endothelin 1 | EDN1 |
| 6496-60 | DLK1:ECD | DLK1 |
| 6504-65 | Lysyl oxidase-like prot | LOXL2 |
| 6518-85 | ghrelin | GHRL |
| 6520-87 | MGP | MGP |
| 6544-33 | NELL1 | NELL1 |
| 6545-58 | PRIO | PRNP |
| 6558-5 | COL10 | COLEC10 |
| 6563-78 | HSP 70 | HSPA1A |
| 6577-64 | LAMA4 | LAMA4 |
| 6584-1 | SRCH | HRC |
| 6593-5 | GALT3 | GALNT3 |
| 6599-5 | APCD1 | APCDD1 |
| 6605-17 | IGFALS | IGFALS |
| 6622-90 | APEL | APLN |
| 6649-51 | NET1 | NTN1 |
| 6895-1 | TR:CD | TFRC |
| 6909-40 | MGAT2 | MGAT2 |
| 6932-42 | ITA5 | ITGA5 |
| 6935-123 | BAMBI:CD | BAMBI |
| 6941-11 | SUMF1 | SUMF1 |
| 6948-82 | FIG4 | FIG4 |
| 6963-82 | SELPL:CD | SELPLG |
| 6973-111 | IGF-II:Pro-form | IGF2 |
| 7015-8 | LIRB5 | LILRB5 |
| 7045-4 | BNIP3 | BNIP3 |
| 7050-5 | NEGR1 | NEGR1 |
| 7100-31 | CD2 | CD2 |
| 7124-18 | IL-21 | IL21 |
| 7127-3 | Apo A-II | APOA2 |
| 7131-8 | CETP | CETP |
| 7140-1 | ELA2A | CELA2A |
| 7142-5 | CBPN | CPN1 |
| 7161-25 | G6PE | H6PD |
| 7174-15 | CD38 | CD38 |
| 7178-59 | DEPP | DEPP1 |
| 7181-17 | VAPB | VAPB |
| 7182-1 | carboxylesterase, liver | CES1 |
| 7186-111 | STX3 | STX3 |
| 7187-3 | PPM1L | PPM1L |
| 7191-32 | SLMAP | SLMAP |
| 7194-36 | NPTN | NPTN |
| 7201-5 | ISCU | ISCU |
| 7206-20 | F16P1 | FBP1 |
| 7247-1 | GORAB | GORAB |
| 7262-191 | CHSTE | CHST14 |
| 7624-19 | ANK2 | ANK2 |

| | | |
|---|---|---|
| 7628-40 | CREL1 | CRELD1 |
| 7655-11 | N-terminal pro-BNP | NPPB |
| 7660-21 | Tropomyosin 4 | TPM4 |
| 7735-17 | PEDF | SERPINF1 |
| 7747-47 | NDUBB | NDUFB11 |
| 7748-11 | NDUV2 | NDUFV2 |
| 7754-11 | JAG1:CD | JAG1 |
| 7761-125 | CHKB | CHKB |
| 7770-25 | NFU1 | NFU1 |
| 7784-1 | Kininogen, HMW | KNG1 |
| 7788-1 | CF6 | ATP5PF |
| 7789-182 | PRDX4 | PRDX4 |
| 7790-21 | LIF | LIF |
| 7806-33 | B4GT7 | B4GALT7 |
| 7849-3 | Glutaminyl cyclase | QPCT |
| 7850-1 | COX42 | COX4I2 |
| 7853-19 | SCO1 | SCO1 |
| 7857-22 | NEUT | NTS |
| 7870-8 | CNDP1 | CNDP1 |
| 7875-86 | PLEK | PLEK |
| 7887-57 | COX5B | COX5B |
| 7888-58 | CCD56 | COA3 |
| 7893-19 | OLR1 | OLR1 |
| 7895-108 | MRAP | MRAP |
| 7905-30 | HPT | HP |
| 7911-29 | SCN4B | SCN4B |
| 7922-5 | Adrenomedullin | ADM |
| 7924-7 | CD3G | CD3G |
| 7953-20 | SLAF1 | SLAMF1 |
| 7981-230 | B3GT6 | B3GALT6 |
| 8005-1 | MXRA7 | MXRA7 |
| 8009-121 | SURF1 | SURF1 |
| 8017-23 | FXRD1 | FOXRED1 |
| 8036-75 | FSHB | FSHB |
| 8038-41 | NSDHL | NSDHL |
| 8043-153 | COMP | COMP |
| 8051-10 | AGGF1 | AGGF1 |
| 8055-33 | IDD | DGCR2 |
| 8059-1 | Tpo | THPO |
| 8067-21 | KCNE3 | KCNE3 |
| 8068-43 | K1161:ECD | MYORG |
| 8070-88 | DLG4 | DLG4 |
| 8074-32 | TMM70 | TMEM70 |
| 8086-49 | ITM2B | ITM2B |
| 8089-173 | NR4A1 | NR4A1 |
| 8091-16 | MASP3:Heavy | MASP1 |
| 8093-13 | K1161:CD | MYORG |

| | | |
|---|---|---|
| 8094-20 | CISD2 | CISD2 |
| 8099-42 | SPON2 | SPON2 |
| 8100-15 | ADM2 | ADM2 |
| 8231-122 | VEGF sR1 | FLT1 |
| 8243-55 | TATI | SPINK1 |
| 8246-9 | PCSK9 | PCSK9 |
| 8250-2 | PTPRJ | PTPRJ |
| 8265-225 | LAP2B | TMPO |
| 8273-84 | IL31R | IL31RA |
| 8285-64 | PACA | ADCYAP1 |
| 8304-50 | OPG | TNFRSF11B |
| 8309-12 | HYAL1 | HYAL1 |
| 8314-71 | GNS | GNS |
| 8353-15 | SCN2B | SCN2B |
| 8356-88 | NEU1 | OXT |
| 8358-30 | Peroxiredoxin-3 | PRDX3 |
| 8364-74 | UST | UST |
| 8368-102 | TNF sR-II | TNFRSF1B |
| 8376-25 | LSHB | LHB |
| 8380-244 | DLK1:CD | DLK1 |
| 8403-18 | Fatty acid synthase | FASN |
| 8406-17 | IGF-I | IGF1 |
| 8429-16 | AXIN2 | AXIN2 |
| 8446-4 | PACAP-27 | ADCYAP1 |
| 8458-16 | a-Synuclein | SNCA |
| 8462-18 | HGH | GH1 |
| 8470-213 | RNase H1 | RNASEH1 |
| 8480-29 | FBLN3 | EFEMP1 |
| 8484-24 | Leptin | LEP |
| 8529-1 | TRAIL R2:Death | TNFRSF10B |
| 8587-21 | ISK52 | SPINK14 |
| 8644-46 | Cathepsin H | CTSH |
| 8645-257 | PEX14:C-term | PEX14 |
| 8687-26 | T106B | TMEM106B |
| 8690-25 | CAV3 | CAV3 |
| 8750-46 | Vinculin | VCL |
| 8756-41 | KCE1L:N-term | KCNE5 |
| 8767-44 | Endothelin-converting | ECE1 |
| 8772-5 | EFNB2:CD | EFNB2 |
| 8790-6 | Arylsulfatase A | ARSA |
| 8795-48 | TR:ECD | TFRC |
| 8810-26 | HEPACAM | HEPACAM |
| 8811-24 | BAMBI:ECD | BAMBI |
| 8834-58 | Calnexin | CANX |
| 8854-59 | RIR2B | RRM2B |
| 8863-3 | P85A | PIK3R1 |
| 8874-53 | CLN5:LD | CLN5 |

| | | |
|---|---|---|
| 8893-29 | PARP:region 1 | PARP1 |
| 8908-14 | KCE1L:CD | KCNE5 |
| 8916-32 | STIM1:CD | STIM1 |
| 8924-55 | CALY | CALY |
| 8932-1 | ENTP6 | ENTPD6 |
| 8935-22 | MRP6 | ABCC6 |
| 8946-38 | NR1H4 | NR1H4 |
| 8952-65 | G-CSF | CSF3 |
| 8955-60 | GL8D1 | GLT8D1 |
| 8985-13 | SRAC1 | SERAC1 |
| 8986-2 | SERA | PHGDH |
| 9000-177 | SLC31 | SLC3A1 |
| 9017-58 | LPH | LCT |
| 9021-1 | TIM-1 | HAVCR1 |
| 9076-25 | PENK | PENK |
| 9125-23 | MASP3:Sushi 1 and Su | MASP1 |
| 9171-11 | CSRP3 | CSRP3 |
| 9173-21 | PGM1 | PGM1 |
| 9176-3 | MUC1:region 2 | MUC1 |
| 9178-30 | NEUREGULIN-1 | NRG1 |
| 9185-15 | TFF1 | TFF1 |
| 9188-119 | MIG | CXCL9 |
| 9197-4 | LEG9 | LGALS9 |
| 9204-33 | Corticotropin-lipotropi | POMC |
| 9213-24 | FTCD | FTCD |
| 9237-54 | Lysosomal acid phosph | ACP2 |
| 9244-27 | PPT1 | PPT1 |
| 9253-52 | BGAT | ABO |
| 9257-14 | TACO1 | TACO1 |
| 9259-35 | DR6 | TNFRSF21 |
| 9269-7 | Biotinidase | BTD |
| 9271-101 | STIM1:ECD | STIM1 |
| 9283-8 | CD44 | CD44 |
| 9287-6 | SPRE | SPR |
| 9337-43 | TKN1 | TAC1 |
| 9340-17 | FKB14 | FKBP14 |
| 9341-1 | PDGFD | PDGFD |
| 9343-16 | IL-2 sRb | IL2RB |
| 9357-4 | CREG1 | CREG1 |
| 9366-54 | IL-21 sR | IL21R |
| 9377-25 | SCF | KITLG |
| 9385-4 | GAA | GAA |
| 9388-18 | MCEE | MCEE |
| 9412-52 | Activin RIIB | ACVR2B |
| 9436-2 | ID-1 | ID1 |
| 9443-137 | cathepsin K | CTSK |
| 9444-70 | DGCR6 | DGCR6 |

| | | |
|---|---|---|
| 9451-20 | Uromodulin | UMOD |
| 9457-3 | CAV2 | CAV2 |
| 9459-7 | Fas, soluble | FAS |
| 9484-75 | Desmoglein-2 | DSG2 |
| 9504-19 | RB27A | RAB27A |
| 9522-3 | AIF | AIFM1 |
| 9543-131 | COG8 | COG8 |
| 9576-58 | MNX1 | MNX1 |
| 9739-4 | MSH2 | MSH2 |
| 9750-7 | S100A4 | S100A4 |
| 9756-6 | TAGL | TAGLN |
| 9759-13 | INDO | IDO1 |
| 9786-310 | IEX1 | IER3 |
| 9789-52 | Nesprin-2 | SYNE2 |
| 9796-4 | CEL | CEL |
| 9800-20 | NDUB8 | NDUFB8 |
| 9802-27 | PLPL2 | PNPLA2 |
| 9823-2 | DYR | DHFR |
| 9830-109 | VAV3 | VAV3 |
| 9832-33 | HGD | HGD |
| 9834-62 | ADH1B | ADH1B |
| 9837-60 | NAD(P)H dehydrogena | NQO1 |
| 9838-4 | SMAD1 | SMAD1 |
| 9842-2 | b-Catenin | CTNNB1 |
| 9844-138 | ACTN2 | ACTN2 |
| 9845-33 | PARK7 | PARK7 |
| 9846-32 | Rho-GDI beta | ARHGDIB |
| 9849-13 | APEX1 | APEX1 |
| 9859-180 | SSAT-1 | SAT1 |
| 9867-23 | F16P2 | FBP2 |
| 9869-28 | NF-kappa-B p105 | NFKB1 |
| 9873-17 | ERRalpha | ESRRA |
| 9874-28 | p15-INK4b | CDKN2B |
| 9877-28 | CRKL | CRKL |
| 9878-3 | SULT 1E | SULT1E1 |
| 9880-33 | T23O | TDO2 |
| 9883-29 | Glyoxalase I | GLO1 |
| 9886-28 | XRCC4 | XRCC4 |
| 9895-77 | XPF | ERCC4 |
| 9897-9 | MCM6 | MCM6 |
| 9900-36 | NFH | NEFH |
| 9923-6 | SOS1 | SOS1 |
| 9937-7 | CXA1 | GJA1 |
| 9952-57 | TFAM | TFAM |

**Table**. Demographic and clinical characteristics of the hypertensive Taiwanese cohort

| Variable | Total (n = 5713) | Q1 (n = 1430) | Q2 (n =1428) | Q3 (n =1427) | Q4 (n =1428) |
|---|---|---|---|---|---|
| Age, year | 59.9 ± 15.4 | 62.8 ± 16.0 | 60.8 ± 15.7 | 59.2 ± 15.0 | 56.9 ± 14.4 |
| Age >65 years | 2207 (38.6) | 659 (46.1) | 598 (41.9) | 520 (36.4) | 430 (30.1) |
| Male sex | 2710 (47.4) | 715 (50.0) | 683 (47.8) | 655 (45.9) | 657 (46.0) |
| Body weight, kg | 66.8 ± 15.0 | 65.0 ± 15.3 | 67.1 ± 15.2 | 67.4 ± 14.9 | 67.6 ± 14.6 |
| Body height, cm | 161.9 ± 8.9 | 161.2 ± 9.0 | 162.3 ± 8.9 | 162.0 ± 8.8 | 162.3 ± 8.7 |
| Body mass index, kg/m2 | 25.3 ± 4.5 | 24.8 ± 4.7 | 25.3 ± 4.6 | 25.5 ± 4.5 | 25.5 ± 4.3 |
| **Biochemical studies** | | | | | |
| Plasma renin activity, ng/mL/h | 0.3 [0.1, 0.6] | 0.3 [0.2, 0.6] | 0.3 [0.2, 0.6] | 0.3 [0.1, 0.6] | 0.3 [0.1, 0.6] |
| Plasma aldosterone concentration, ng/dL | 20.6 [12.3, 32.9] | 8.1 [5.1, 10.6] | 15.9 [14.0, 18.1] | 26.3 [23.4, 29.5] | 44.9 [37.2, 61.2] |
| Aldosterone-to-renin ratio, ng/dL per ng/mL/h | 66.6 [30.7, 173.2] | 21.7 [11.6, 50.1] | 51.5 [27.0, 97.5] | 83.4 [42.5, 189.1] | 203.7 [89.8, 529.3] |
| Hemoglobin, g/dL | 12.9 ± 2.3 | 12.3 ± 2.4 | 12.9 ± 2.4 | 13.2 ± 2.2 | 13.3 ± 2.0 |
| Serum creatinine, mg/dL | 1.2 ± 1.0 | 1.2 ± 1.1 | 1.2 ± 1.1 | 1.1 ± 0.9 | 1.2 ± 0.9 |
| eGFR, mL/min/1.73m2 | 81.4 ± 33.3 | 84.0 ± 37.2 | 79.8 ± 32.9 | 80.4 ± 30.6 | 81.5 ± 32.1 |
| Sodium, mEq/L | 138.8 ± 3.9 | 138.2 ± 4.3 | 138.7 ± 3.8 | 138.8 ± 3.7 | 139.5 ± 3.7 |
| Potassium, mEq/L | 4.0 ± 0.6 | 3.9 ± 0.6 | 4.0 ± 0.6 | 4.0 ± 0.6 | 3.9 ± 0.6 |
| Fasting glucose, mg/dL | 108.5 ± 31.4 | 113.7 ± 37.0 | 109.9 ± 33.8 | 106.3 ± 27.1 | 104.4 ± 26.2 |
| ALT, U/L | 25.9 ± 21.7 | 25.9 ± 21.7 | 26.6 ± 25.6 | 25.8 ± 21.5 | 24.9 ± 19.0 |
| **Comorbidity** | | | | | |
| Diabetes mellitus | 1124 (19.7) | 372 (26.0) | 292 (20.4) | 251 (17.6) | 209 (14.6) |

| Variable | Total (n = 5713) | Q1 (n = 1430) | Q2 (n =1428) | Q3 (n =1427) | Q4 (n =1428) |
|---|---|---|---|---|---|
| Hyperlipidemia | 1206 (21.1) | 336 (23.5) | 318 (22.3) | 291 (20.4) | 261 (18.3) |
| Coronary artery disease | 610 (10.7) | 168 (11.7) | 137 (9.6) | 157 (11.0) | 148 (10.4) |
| Atrial fibrillation | 174 (3.0) | 49 (3.4) | 55 (3.9) | 43 (3.0) | 27 (1.9) |
| Obstructive sleep apnea | 193 (3.4) | 47 (3.3) | 59 (4.1) | 45 (3.2) | 42 (2.9) |
| Chronic kidney disease | 822 (14.4) | 259 (18.1) | 222 (15.5) | 179 (12.5) | 162 (11.3) |
| Heart failure | 215 (3.8) | 82 (5.7) | 66 (4.6) | 44 (3.1) | 23 (1.6) |
| Acute myocardial infarction | 66 (1.2) | 27 (1.9) | 14 (1.0) | 17 (1.2) | 8 (0.6) |
| Intracranial hemorrhage | 221 (3.9) | 80 (5.6) | 64 (4.5) | 46 (3.2) | 31 (2.2) |
| Ischemic stroke | 517 (9.0) | 190 (13.3) | 138 (9.7) | 117 (8.2) | 72 (5.0) |
| **Anti-hypertensive medications at baseline** | | | | | |
| RASi | 2958 (51.8) | 720 (50.3) | 749 (52.5) | 747 (52.3) | 742 (52.0) |
| Beta-blocker | 2341 (41.0) | 581 (40.6) | 607 (42.5) | 586 (41.1) | 567 (39.7) |
| Alpha-blocker | 1344 (23.5) | 259 (18.1) | 295 (20.7) | 356 (24.9) | 434 (30.4) |
| Vasodilator | 439 (7.7) | 118 (8.3) | 119 (8.3) | 109 (7.6) | 93 (6.5) |
| Thiazide | 577 (10.1) | 108 (7.6) | 141 (9.9) | 180 (12.6) | 148 (10.4) |
| Calcium channel blocker | 3535 (61.9) | 795 (55.6) | 861 (60.3) | 933 (65.4) | 946 (66.2) |
| Others (Clonidine, Minoxidil) | 238 (4.2) | 59 (4.1) | 53 (3.7) | 61 (4.3) | 65 (4.6) |
| MRA | 277 (4.8) | 66 (4.6) | 47 (3.3) | 57 (4.0) | 107 (7.5) |
| Number of medications | 2.1 ± 1.6 | 2.0 ± 1.7 | 2.1 ± 1.6 | 2.2 ± 1.6 | 2.2 ± 1.6 |

Abbreviation: ALT, alanine aminotransferase; eGFR, estimated glomerular filtration rate; MRA, mineralocorticoid receptor antagonist; RASi, renin-angiotensin system inhibitors.

Data are presented as number (percentage), mean ± standard deviation or median [25$^{th}$, 75$^{th}$ percentiles].
